# FORECAST-RBD: Forecasting Phenoconversion Risks and Its Clinical Phenotype in Patients with Isolated RBD Using Machine Learning and Explainable AI

**DOI:** 10.1101/2024.06.02.24308240

**Authors:** Yong Woo Shin, Jung-Ick Byun, Han-Joon Kim, Ki-Young Jung

**Affiliations:** Department of Neurology, Inha University Hospital, Incheon, South Korea; Department of Neurology, Kyung Hee University Hospital at Gangdong, Seoul, South Korea; Department of Neurology, Seoul National University Hospital, Seoul National University College of Medicine, Seoul, South Korea

## Abstract

**Background and Objectives:** Isolated Rapid Eye Movement (REM) sleep behavior disorder (iRBD) is a sleep disorder associated with neurodegenerative diseases such as Parkinson’s disease and dementia with Lewy bodies. Predicting which iRBD patients will phenoconvert to neurodegenerative diseases is crucial for prognosis and management. The objective of this study is to develop a machine learning model using clinical markers to predict phenoconversion in patients with RBD.

**Methods:** Analyzing a cohort of 178 iRBD patients over a median follow-up period of 3.6 years, during which 30 patients converted to neurodegenerative conditions, we leveraged an comprehensive dataset encompassing demographics, medication history, cognitive assessments, sleep quality, autonomic symptoms, and parkinsonian signs. We explored a variety of feature selection methods and survival models. Additionally, separate models to predict the subtype of photoconversion—whether motor-first or cognition-first—were developed.

**Results:** The extreme gradient boosting survival embeddings-Kaplan neighbors (XGBSE-KN) model demonstrated the best performance, achieving a concordance index of 0.823 and integrated Brier score of 0.123 on 10-fold cross-validation. Explainable AI methods provided insights into prediction rationales and key risk factors including age, RBDQ-KR factor 2 (behavioral factors), weight, antidepressant, coffee use, and UPDRS III excluding tremor score. For subtype classification, the RandomForestClassifier utilizing three features (PSQI-TST, MoCA, and age), emerged as the most effective, achieving a Matthews Correlation Coefficient of 0.697 in 100 repeated 5-fold cross-validations. These models have been deployed on a server for physician access.

**Discussion:** These models can aid prognosis and enable personalized management in RBD patients, potentially improving patient care and outcomes. While these findings are promising, further external validation of the models is necessary to confirm their efficacy and reliability in clinical settings. Future research should focus on incorporating additional biomarkers and exploring the models’ performance in larger, diverse cohorts.

## Introduction

Isolated rapid eye movement (REM) behavior disorder (iRBD) is one of the early sign of alpha-synuclein-mediated neurodegenerative diseases.^1–4^ The progression from iRBD to neurodegenerative diseases such as Parkinson’s disease and dementia with Lewy bodies varies from months to decades, with an annual conversion rate of about 6% and around 75% within 12 years of diagnosis.^5,6^ Identifying those at higher risk of such neurodegenerative diseases is essential not only for advancing neuroprotective trials but also for patient life planning and management, offering opportunities for early intervention and personalized patient management.

Prior research has highlighted several risk factors associated with the phenoconversion of patients with iRBD to neurodegenerative diseases. Longitudinal studies have identified factors that increase the risk, including advanced age, the presence of hyposmia, abnormalities in color vision, mild signs of parkinsonism, slight cognitive decline, disturbances in autonomic functions, impairment in nigrostriatal dopaminergic pathways, and the extent of REM sleep without atonia (RSWA) loss. These insights have been crucial for identifying individuals with iRBD who are at an elevated risk of developing neurodegenerative conditions. ^5,6^ However, these findings are often limited in their application in clinical settings due to resource constraints or the rarity of certain features. Additionally, while previous studies have identified risk factors associated with phenoconversion, they have not successfully translated these findings into personalized, actionable prediction models for clinical use.

Prognostic counseling in iRBD patients is crucial for managing future health risks, and enabling early interventions, while also providing necessary psychological support. A previous study showed that most of the patients expressed a strong preference for detailed prognostic information.^7^ However, it is often challenging to discuss prognostication, given the uncertainty in accurate personalized prediction, and the lack of a standardized approach.^8^ This underscores the urgent need for tools that can support prognostication.

To address these issues, our study leverages machine learning for predicting phenoconversion time and subtype in iRBD using clinical markers, thereby providing personalized, actionable prediction models. These models, developed to overcome the limitations of traditional survival analysis methods like the Cox Proportional Hazards model (CoxPH), offer an advanced approach to clinical decision-making. CoxPH, while theoretically solid, is limited by its reliance on linear feature interactions and its inadequacy in handling multicollinearity or large-scale, high-dimensional datasets. The resultant models not only aid in prognosis but also enhance clinical decision-making, providing insights into disease progression tailored to individual patient profiles.

This study aims to develop and implement machine learning-based models for predicting both the timing and subtype of phenoconversion in patients with iRBD. By utilizing clinical variables, we aim to bridge the gap between research and practice, offering a web-based tool for clinicians to facilitate early identification of high-risk patients and enable timely, personalized medical care.

## Methods

### Study Population and Data Collection

Patients were recruited from the rapid eye movement (REM) sleep behavior disorder (RBD) registry at the Seoul National University Hospital between April 2016 and May 2022. Participants underwent diagnostic overnight video-polysomnography (vPSG) and were enrolled upon meeting International Classification of Sleep Disorders, 3rd edition (ICSD-3) criteria for isolated RBD. At recruitment, two neurologists specializing in sleep medicine (JK) and movement disorders (KH) performed comprehensive evaluations to exclude secondary causes of RBD or comorbid neurodegenerative diseases, or major medical illnesses.

Individuals with any preexisting neurodegenerative diseases like Parkinson’s disease, dementia with Lewy bodies, or multiple system atrophy were excluded from the study. In addition, the study’s exclusion criteria also included those with a history of neurological disorders like epilepsy or stroke, past psychiatric illnesses, head trauma, and recent use of medications that could affect sleep or motor functions. Severe obstructive sleep apnea, defined as having an apnea-hypopnea index of 30 or higher on baseline video-polysomnography (vPSG), was another key exclusion factor. Moreover, individuals with serious medical comorbidities were not considered for enrollment. Participants who met all the eligibility criteria provided written informed consent before being included in the study, which was approved by the Institutional Review Board of Seoul National University Hospital (IRB No.: 1708-169-883).

### Clinical evaluation

Various demographic, medical history, and clinical evaluation data were collected from participants. Demographic variables included age, sex, height, weight, and body mass index (BMI). Past medical history encompassed medical conditions, psychiatric illness, antidepressant use, alcohol use, smoking, pesticide exposure, solvent exposure, and first-degree relatives with Parkinson’s disease. Clinical assessments evaluated olfactory loss, prior injury, daily caffeine and coffee consumption, and years of education. We evaluated olfactory loss using the Korean Version of the Sniffin’ Sticks test (KVSS). ^9^ RBD symptom frequency and severity were measured with the REM Sleep Behavior Disorder Questionnaire (RBDQ-KR).^10^ Cognitive functions were evaluated using the Mini-Mental State Exam (MMSE), Montreal Cognitive Assessment (MoCA).^11,12^ Sleep quality and daytime sleepiness assessments included Epworth Sleepiness Scale (ESS), Insomnia Severity Index (ISI), and Pittsburgh Sleep Quality Index (PSQI).^13–15^ We evaluated the mental health of the participants through the Geriatric Depression Scale (GDS).^16^ Autonomic dysfunction was quantified by the SCOPA-AUT scale including GI, urinary, cardiovascular, and sexual domains.^17^ Motor performance was graded with the Movement Disorders Society-Unified Parkinson’s Disease Rating Scale (MDS-UPDRS) Part III.^18^ Phenoconversion in iRBD patients was assessed every 6 to 12 months by the same two neurologists (JK and KH).^19–21^

### Data Preparation and Imputation for Model Predictors

Variables with greater than 30 percent missing values were excluded from the analysis. For the remaining variables, we employed the Multiple Imputation by Chained Equations (MICE) methodology to impute missing values, using logistic regression for binary variables, polynomial logistic regression for categorical variables, and Predictive Mean Matching (PMM) for continuous variables. Nominal variables were encoded using one-hot encoding or binary encoding. Ordinal variables were converted to numeric values using label encoding. Only baseline measurements were utilized as model predictors.

### Model Development – Prediction of Phenoconversion Time

Four feature selection techniques were applied: univariate filtering, L1 regularization, recursive feature elimination (RFE), and SelectKBest (SKB). RFE and SKB utilized permutation importance based on random survival forest ^22^, and gradient boosting Cox models, with performance measured by concordance index and integrated Brier score (IBS). When the feature set maximizing concordance index differed from the set maximizing IBS, each subset was evaluated separately. The concordance index, or C-index, measures the predictive accuracy of the model, reflecting the proportion of patient pairs whose predicted survival outcomes are correctly ordered by the model. It ranges from 0.5 (no better than chance) to 1.0 (perfect prediction). The integrated Brier score (IBS) quantifies the accuracy of survival probabilities, where a lower score indicates more accurate predictions. The C-index reflects how well the model ranks the survival times, while the IBS measures the calibration and refinement of the survival probabilities. Only the feature sets that maximized these metrics were reported and used.

Survival analysis machine learning models were developed and rigorously evaluated using 10-fold cross-validation to ensure robustness and generalizability. These models included elastic net, accelerated failure time models, random survival forests, survival tree, gradient boosting machines (GBM), and Extreme Gradient Boosting Survival Embeddings (XGBSE). Hyperparameter optimization for each model was conducted using Bayesian optimization. To compare the performance of the developed machine learning models in survival analysis, we adopted the corrected resampled paired t-test method as proposed by Nadeau and Bengio. This method is specifically designed to address the dependencies that may exist in datasets due to repeated measures or resampling techniques. By applying this correction, we ensured that our statistical inferences about the model performances were more robust and reliable.^23^ For multiple testing correction, we incorporated the Benjamini-Hochberg (BH) procedure.

All analyses were conducted using Python 3.10.9, with key libraries including scikit-learn (v.1.1.3), scikit-survival (v.0.19.0), lifelines (v.0.27.4), optuna (v.3.2.0), pandas (v1.5.3), numpy (v1.22.3), and xgboost-survival-embeddings or XGBSE (v0.2.3). Additionally, matplotlib (v3.7.1), and seaborn (v0.11.2) were used for data visualization.

### Model Development – Prediction of Phenoconversion Subtype

Our investigation expanded to include the development of predictive models for the subtype of phenoconversion in iRBD, differentiating between ‘motor-first’ subtypes, such as Parkinson’s Disease (PD) and Multiple System Atrophy (MSA), and the ‘cognition-first’ subtype, exclusively Dementia with Lewy Bodies (DLB), noted for its initial cognitive decline. We applied univariable analysis for initial feature selection to identify predictors differentiating between ‘motor-first’ and ‘cognition-first’ subtypes. To capture non-linear relationships, we utilized the Minimum-redundancy-maximum-relevance (mRMR) feature selection method with Maximal Information Coefficient (MIC) for relevance assessment.^24,25^ The optimal number of features was determined by assessing the Random Forest model’s performance, refining our model to effectively predict RBD subtypes. We developed eleven classifiers using various machine learning algorithms including Random Forest, Support Vector Machines, and Multi-layer Perceptron. To tackle the challenges of the small and imbalanced dataset in our RBD subtype prediction model, we integrated the Synthetic Minority Over-sampling Technique (SMOTE) and the Self-Training method, a semi-supervised learning approach.^26,27^ SMOTE was used to balance the class distribution, while Self-Training leveraged unlabeled data to augment the training dataset, enhancing the overall effectiveness of our predictive models. Censored data were used as unlabeled data in the Self Training. For our subtype prediction model in REM Sleep Behavior Disorder (RBD) with only 30 patients, we used a 100 repeated 5-fold Stratified Cross-Validation due to the small and imbalanced nature of the dataset. This approach was chosen over a 10-fold CV to obtain more stable and reliable estimates by increasing the number of repetitions and ensuring that each data point was adequately represented in both the training and validation sets. For statistical analysis, we applied the correction method by Bouckaert and Frank (2004) and used the R library CorrectR. For multiple testing correction, we incorporated the Benjamini-Hochberg (BH) procedure.

The Matthews Correlation Coefficient (MCC) was chosen as the primary evaluation metric because of its robustness for binary classification, especially with imbalanced class distributions. MCC scores range from −1 to 1, with 1 indicating perfect prediction, 0 representing random guessing, and −1 meaning total disagreement. Along with MCC, several secondary performance metrics were also calculated, including F1 score, precision, recall, and accuracy. Separately, the Receiver Operating Characteristic Area Under the Curve (ROC-AUC) was calculated using a leave-pair-out cross-validation approach. This method was specifically chosen for its ability to minimize bias in the evaluation of the AUC, particularly important in studies with smaller sample sizes.^28^ All analyses were conducted using Python 3.10.9, with key libraries including scikit-learn (v.1.1.3), xgboost (v1.7.5), optuna (v.3.2.0), pandas (v1.5.3), and numpy (v1.22.3). Additionally, matplotlib (v3.7.1), and seaborn (v0.11.2) were used for data visualization.

### Model Explanation and Deployment

Kernel SHAP and time varying SHAP methods provided model explanations by scoring feature impact. For the phenoconversion time prediction model, Kernel SHAP (SHapley Additive exPlanations) was employed to interpret the contributions of individual features. ^29^ Additionally, we incorporated SurvSHAP, a novel method for time-varying feature importance in survival models, which allowed for a more dynamic understanding of how different variables influence the risk of phenoconversion over time.^30^

The generated model was deployed as a web application for facilitating clinician use. An overview of the study design and patient selection are presented in Supplementary Figure 1.

## Results

### Study Population

A total of 417 patients from the sleep center’s RBD registry were approached for the study. Following exclusions, 178 patients were eligible and included for subsequent analysis (Figure 1). Of these 178 participants, 30 developed a neurodegenerative disease during the median follow-up period of 3.6 years. The risk for developing neurodegenerative diseases was 4.81% at two years follow-up, 18.34% at four years and 23.31% at 6 years (Figure 2). Among the patients who phenoconverted, Parkinson’s Disease (PD) was the most prevalent diagnosis, accounting for 50% of cases (15 patients). This was followed by Dementia with Lewy Bodies (DLB), representing 30% of conversions (9 patients), and Multiple System Atrophy-Cerebellar type (MSA-C), comprising 20% (6 patients). The remaining 148 patients did not develop any neurodegenerative disease within the study period and were categorized as still disease-free.

**Figure 1.**
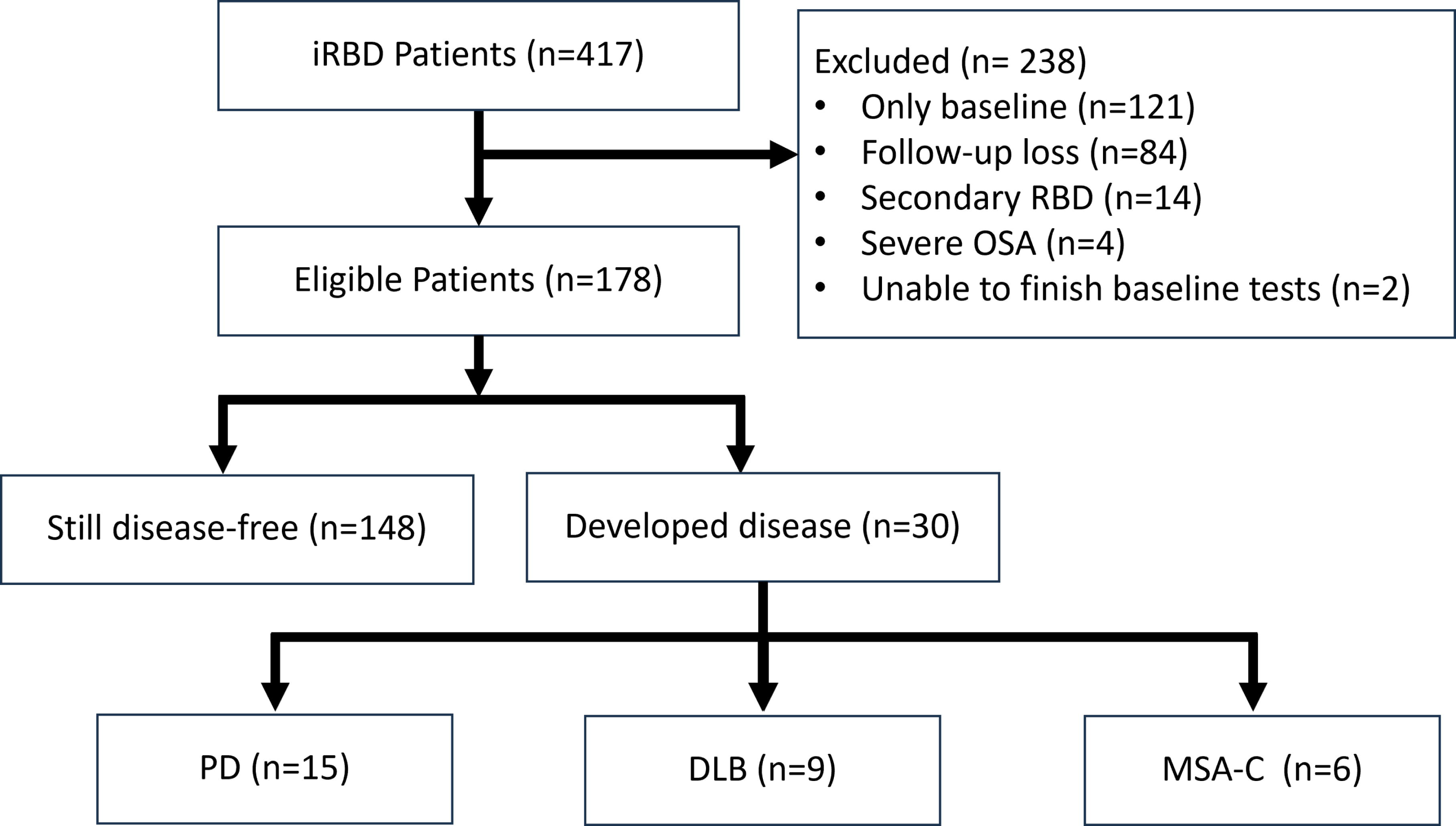
Flow-Chart of Patient Selection

**Figure 2.**
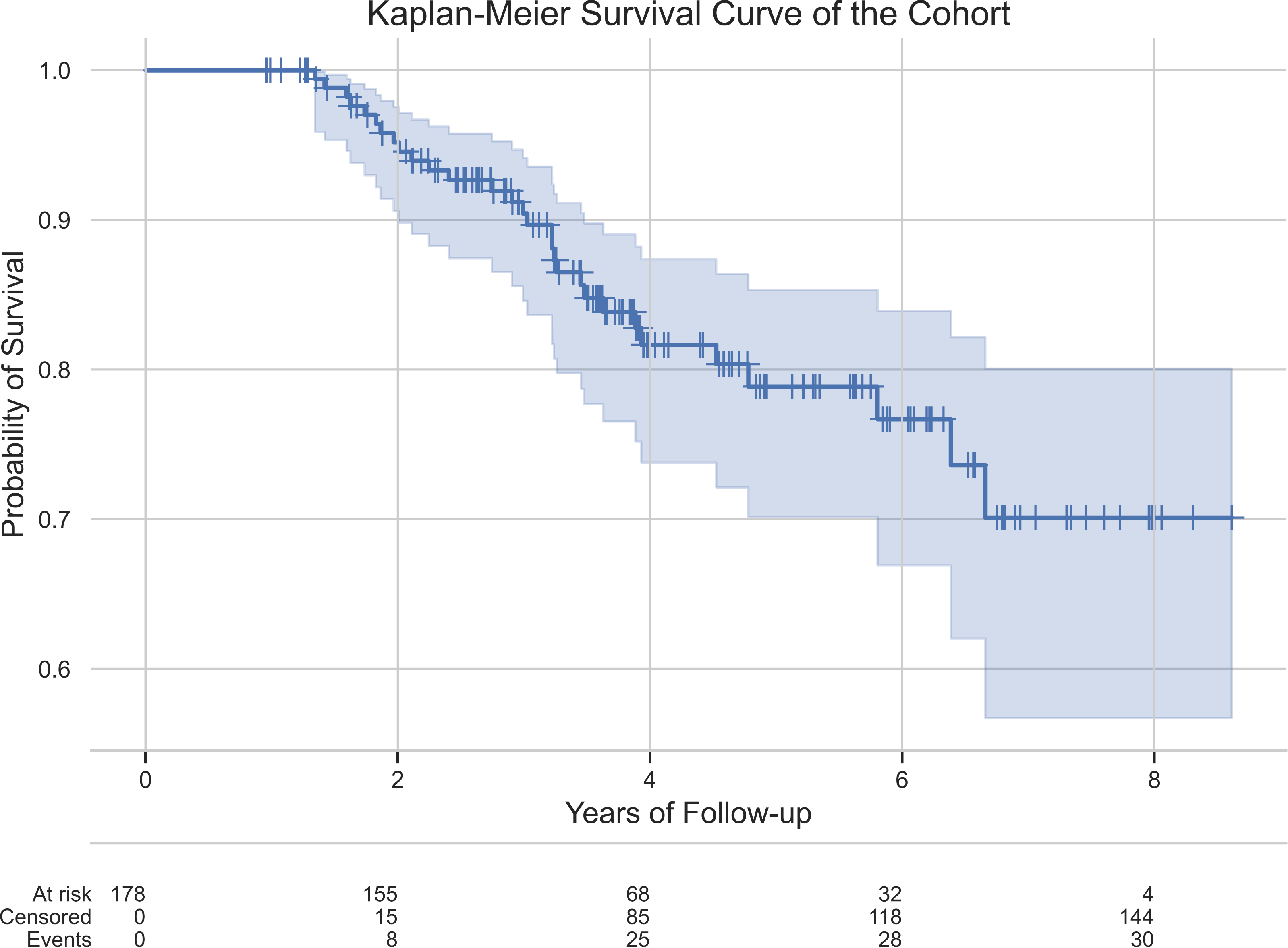
Kaplan-Meier Estimate of the Cohort

### Clinical and Demographic Characteristics

Each prodromal marker was analyzed as a continuous variable or a categorical variable, as appropriate. To assess the risk associated with each factor, hazard ratios (HRs) were calculated using Cox Proportional Hazards (CoxPH) modeling. This approach was employed for both unadjusted and age- and sex-adjusted analyses where applicable. Several factors emerged as significant predictors of phenoconversion. These included age, with an HR of 1.09 (95% CI [1.03-1.15]); use of antidepressants, HR 3.69 (95% CI [1.62-8.44]); solvent exposure, HR 2.73 (95% CI [1.24-5.99]); and caffeine use, HR 0.42 (95% CI [0.20-0.90]). Furthermore, components of the Pittsburgh Sleep Quality Index, specifically PSQI-C3 (sleep duration) and PSQI-C4 (sleep efficiency), yielded HRs of 0.61 (95% CI [0.41-0.92]) and 0.64 (95% CI [0.44-0.92]), respectively. Motor Examination (Part III) of the Movement Disorders Society-Unified Parkinson’s Disease Rating Scale (MDS-UPDRS), excluding tremor scores, also proved significant with an HR of 1.32 (95% CI [1.07-1.63]). The most taken antidepressant was escitalopram (n=11), followed by nortriptyline (n=4), paroxetine (n=2), and agomelatine (n=1).

We analyzed the characteristics of RBD patients based on the subtype of phenoconversion: cognition-first (n=9) and motor-first (n=21). Significant differences were noted between the groups. Age was a distinguishing factor; the cognition-first group was significantly older (cognition-first: 75.0 years versus. motor-first: 67.0 years; p=0.02). Differential performance on cognitive assessments was also evident; those in the cognition-first group scored lower on both MMSE (cognition-first: 26.0 versus motor-first: 28.0; p=0.01), and MoCA (cognition-first: 23.0 versus motor-first: 26.0; p=0.03). Furthermore, an intriguing difference was observed in the Total Sleep Time (TST) component of the Pittsburgh Sleep Quality Index (PSQI), with the cognition-first group experiencing significantly longer sleep durations (cognition-first: 8.4 hours versus motor-first: 7.0 hours; p=0.03). The PSQI-C3 also addresses total sleep time, but there were no significant differences. The lack of a difference is likely due to the fact that the median PSSI-TST for the cog-first group was 8.4 hours and 7.0 for motor-first, with the majority being over 7 hours, and the PSQI-C3 is scored in such a way that total sleep time below 7 hours is associated with higher scores.

### Phenoconversion Time Prediction

The evaluation of phenoconversion prediction models was conducted using four feature selection methods, and 10 models with 10-fold cross validation (Table 3, 4, Supplementary Table 1). In our analysis of phenoconversion time prediction models, the XGBSE-KN model, featuring predictors selected by Recursive Feature Elimination with Random Survival Forest (RFE-RSF), emerged as the most effective, achieving a C-index of 0.823, indicating a high degree of accuracy in predicting the correct sequence of patient phenoconversion events, and an Integrated Brier Score (IBS) of 0.123. (Supplementary Figure 2A, 3A)

**Table 1.**
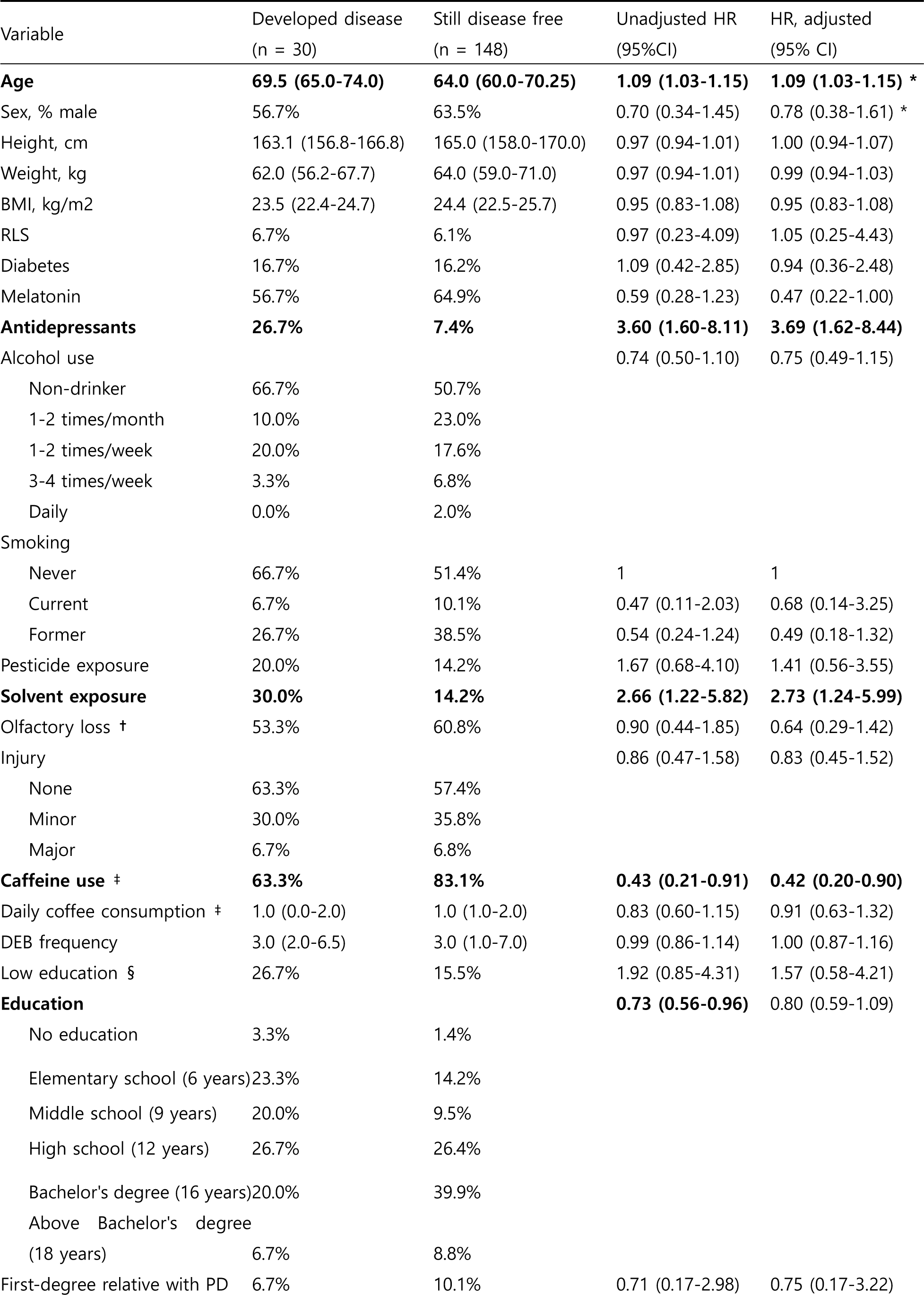
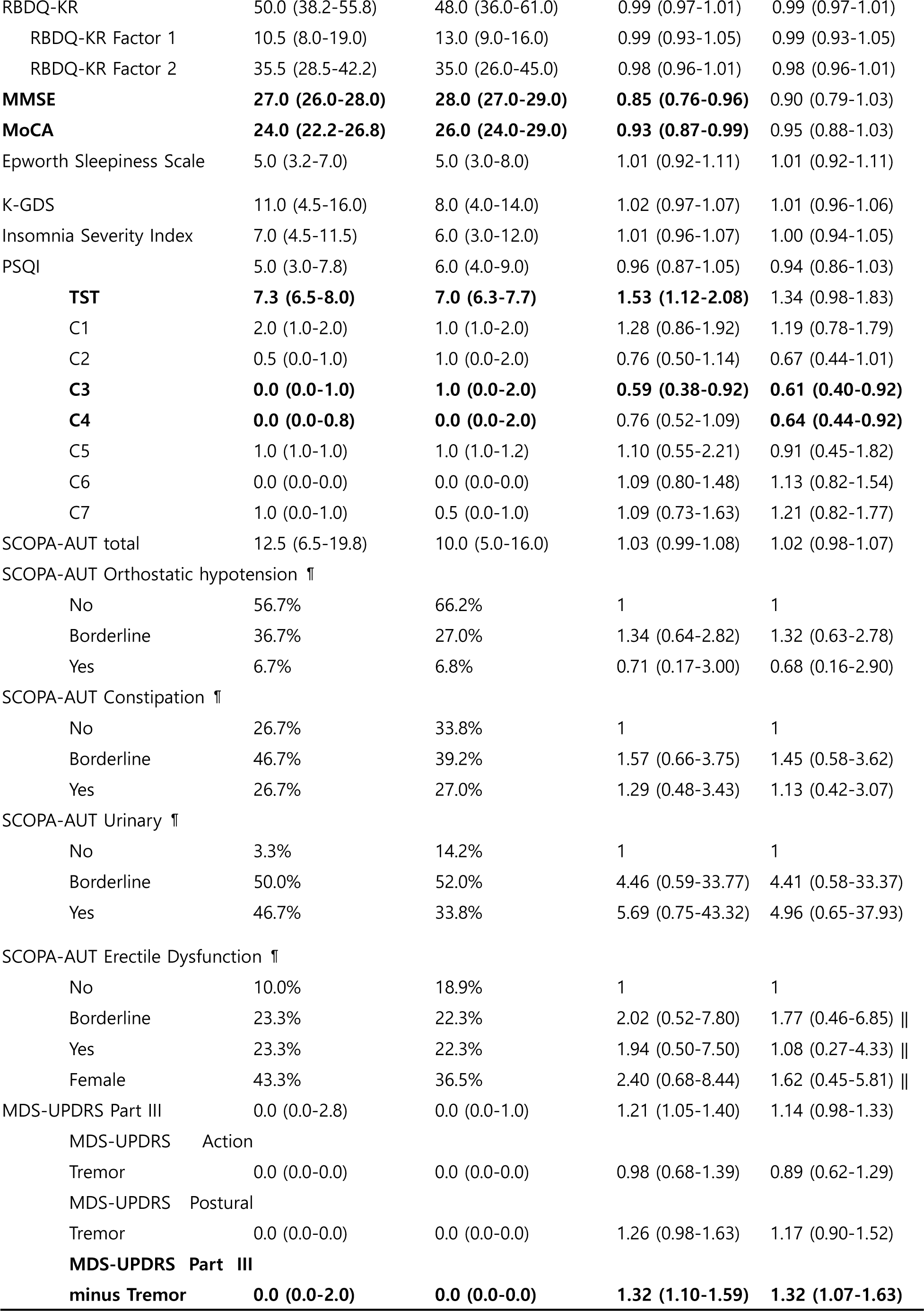
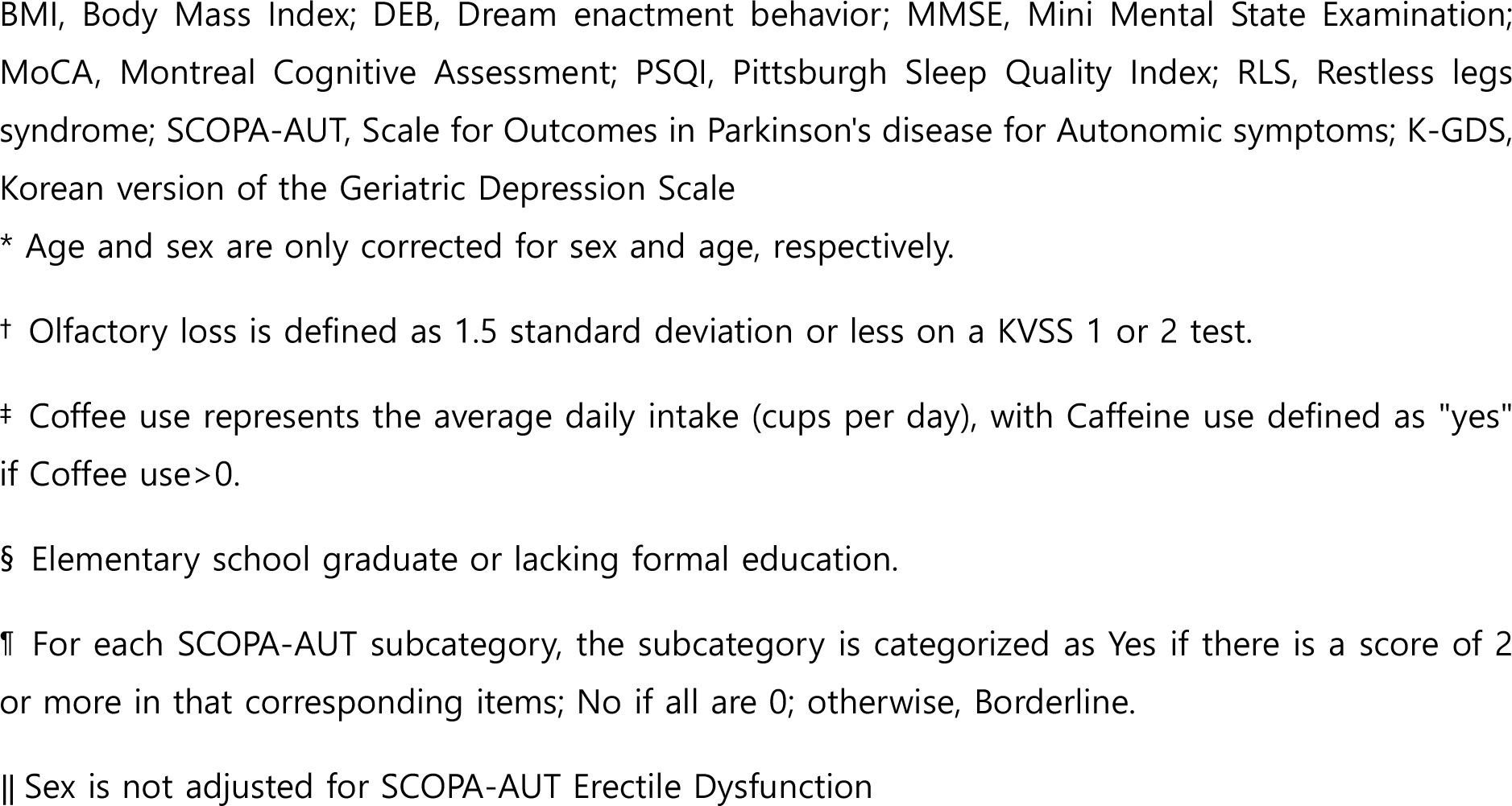
Baseline predictors of neurodegenerative phenoconversion in iRBD.

**Table 2.**
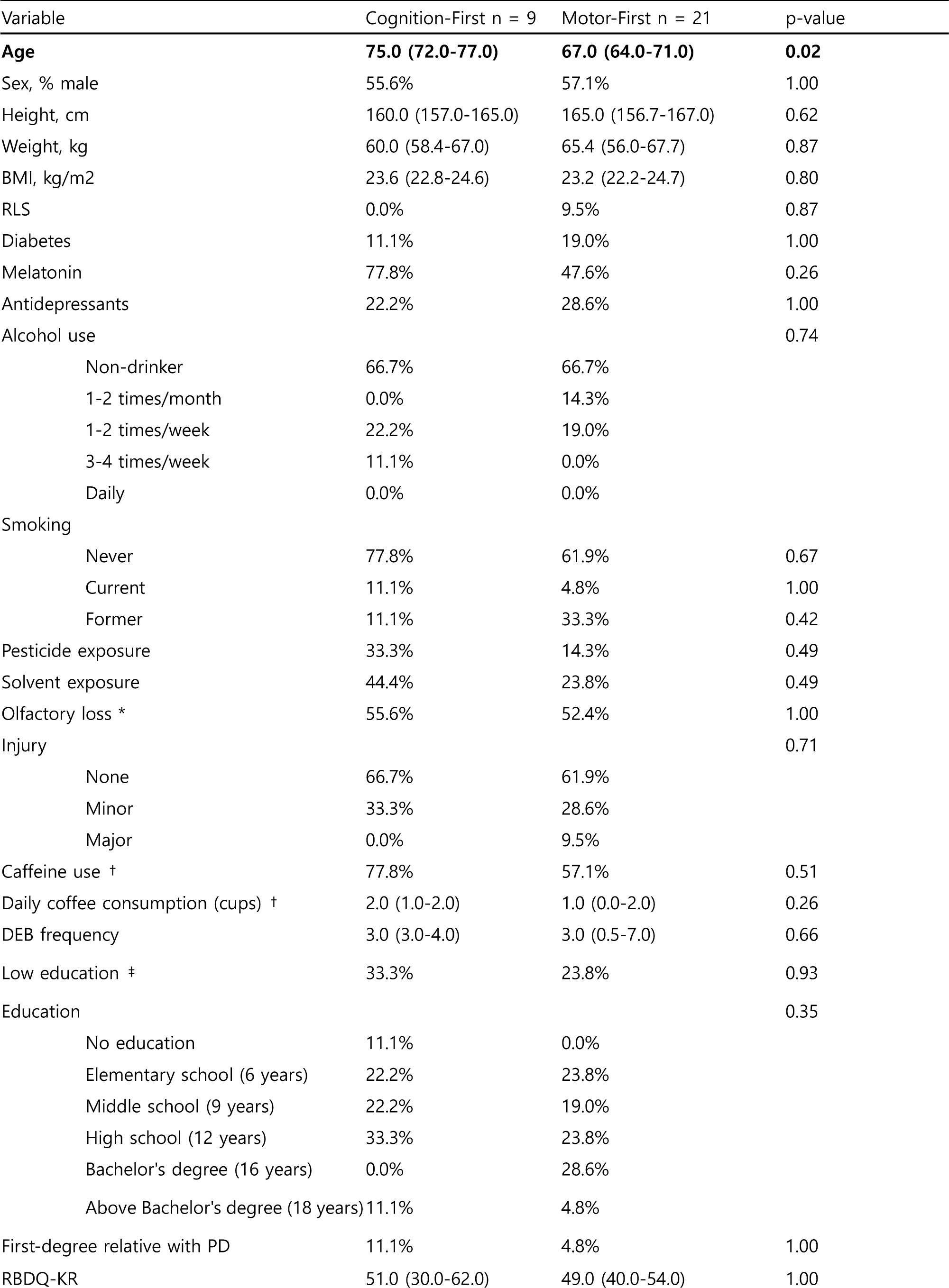
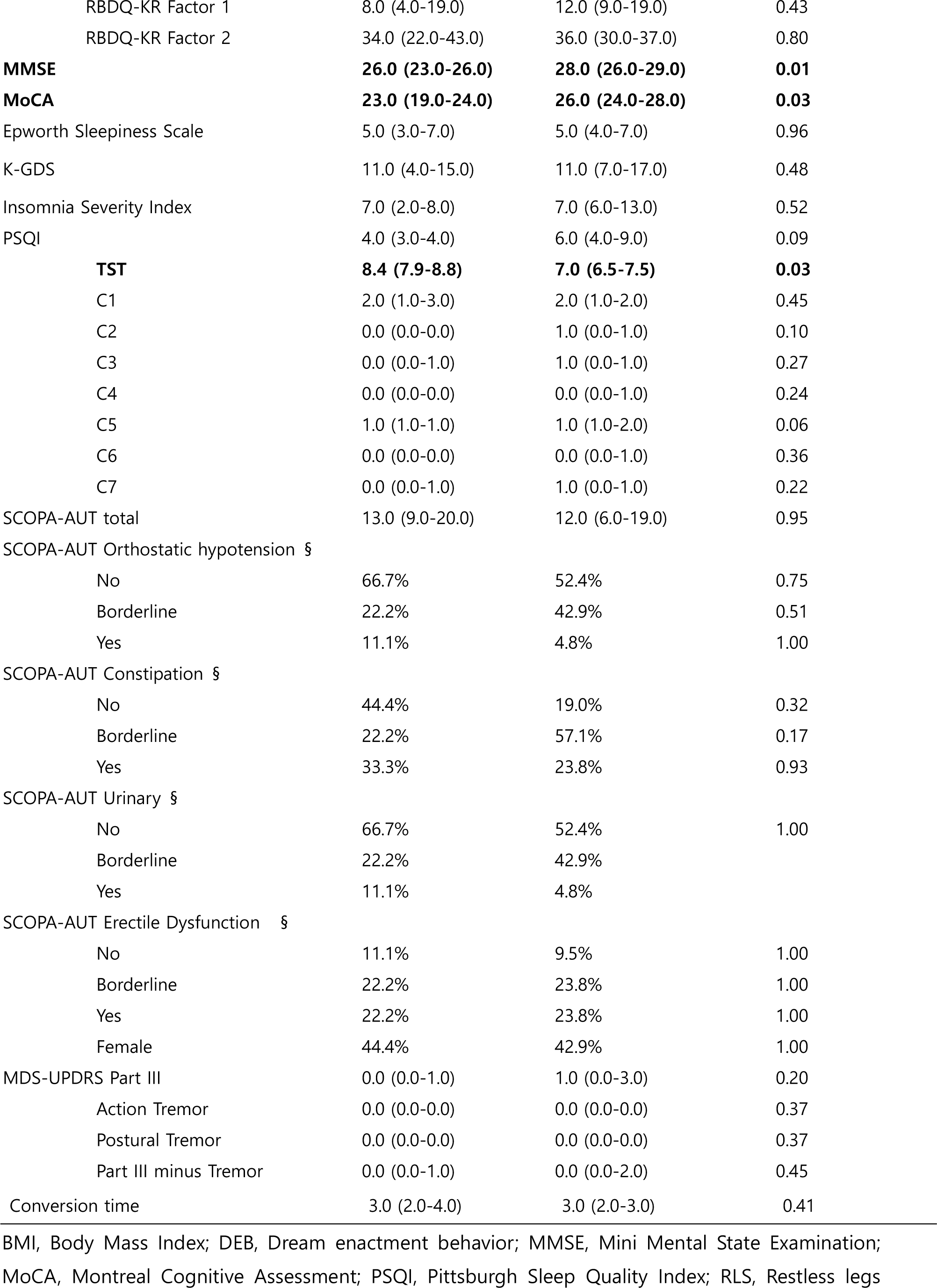
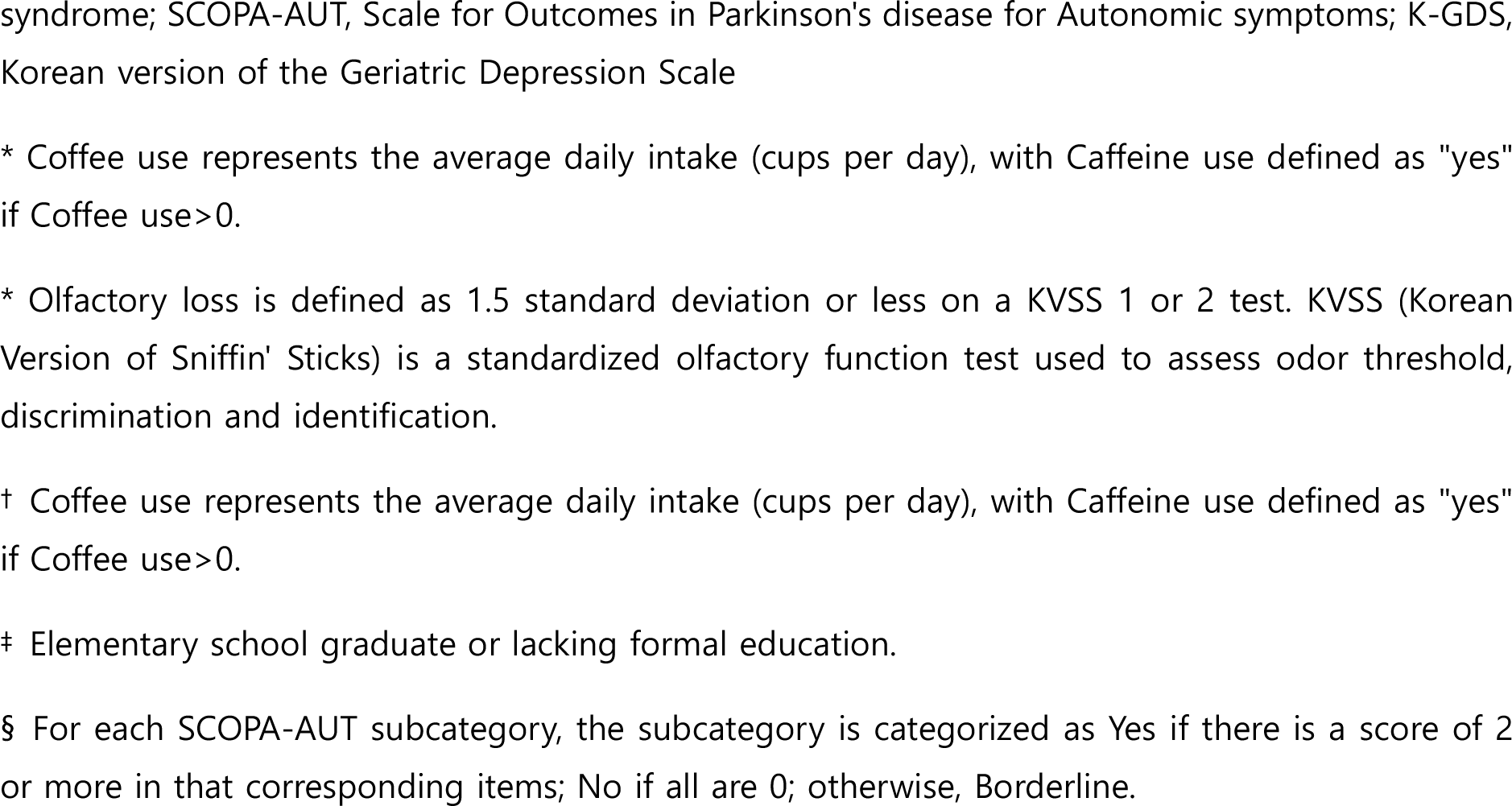
Participant characteristics of cognition- and motor-first subtypes.

| Variable | Cognition-First n = 9 | Motor-First n = 21 | p-value |
| --- | --- | --- | --- |
| **Age** | **75.0 (72.0-77.0)** | **67.0 (64.0-71.0)** | **0.02** |
| Sex, % male | 55.6% | 57.1% | 1.00 |
| Height, cm | 160.0 (157.0-165.0) | 165.0 (156.7-167.0) | 0.62 |
| Weight, kg | 60.0 (58.4-67.0) | 65.4 (56.0-67.7) | 0.87 |
| BMI, kg/m<sup>2</sup> | 23.6 (22.8-24.6) | 23.2 (22.2-24.7) | 0.80 |
| RLS | 0.0% | 9.5% | 0.87 |
| Diabetes | 11.1% | 19.0% | 1.00 |
| Melatonin | 77.8% | 47.6% | 0.26 |
| Antidepressants | 22.2% | 28.6% | 1.00 |
| Alcohol use |  |  | 0.74 |
| Non-drinker | 66.7% | 66.7% |  |
| 1-2 times/month | 0.0% | 14.3% |  |
| 1-2 times/week | 22.2% | 19.0% |  |
| 3-4 times/week | 11.1% | 0.0% |  |
| Daily | 0.0% | 0.0% |  |
| Smoking |  |  |  |
| Never | 77.8% | 61.9% | 0.67 |
| Current | 11.1% | 4.8% | 1.00 |
| Former | 11.1% | 33.3% | 0.42 |
| Pesticide exposure | 33.3% | 14.3% | 0.49 |
| Solvent exposure | 44.4% | 23.8% | 0.49 |
| Olfactory loss * | 55.6% | 52.4% | 1.00 |
| Injury |  |  | 0.71 |
| None | 66.7% | 61.9% |  |
| Minor | 33.3% | 28.6% |  |
| Major | 0.0% | 9.5% |  |
| Caffeine use † | 77.8% | 57.1% | 0.51 |
| Daily coffee consumption (cups) † | 2.0 (1.0-2.0) | 1.0 (0.0-2.0) | 0.26 |
| DEB frequency | 3.0 (3.0-4.0) | 3.0 (0.5-7.0) | 0.66 |
| Low education ‡ | 33.3% | 23.8% | 0.93 |
| Education |  |  | 0.35 |
| No education | 11.1% | 0.0% |  |
| Elementary school (6 years) | 22.2% | 23.8% |  |
| Middle school (9 years) | 22.2% | 19.0% |  |
| High school (12 years) | 33.3% | 23.8% |  |
| Bachelor's degree (16 years) | 0.0% | 28.6% |  |
| Above Bachelor's degree (18 years) | 11.1% | 4.8% |  |
| First-degree relative with PD | 11.1% | 4.8% | 1.00 |
| RBDQ-KR | 51.0 (30.0-62.0) | 49.0 (40.0-54.0) | 1.00 |
| RBDQ-KR Factor 1 | 8.0 (4.0-19.0) | 12.0 (9.0-19.0) | 0.43 |
| RBDQ-KR Factor 2 | 34.0 (22.0-43.0) | 36.0 (30.0-37.0) | 0.80 |
| **MMSE** | **26.0 (23.0-26.0)** | **28.0 (26.0-29.0)** | **0.01** |
| **MoCA** | **23.0 (19.0-24.0)** | **26.0 (24.0-28.0)** | **0.03** |
| Epworth Sleepiness Scale | 5.0 (3.0-7.0) | 5.0 (4.0-7.0) | 0.96 |
| K-GDS | 11.0 (4.0-15.0) | 11.0 (7.0-17.0) | 0.48 |
| Insomnia Severity Index | 7.0 (2.0-8.0) | 7.0 (6.0-13.0) | 0.52 |
| PSQI | 4.0 (3.0-4.0) | 6.0 (4.0-9.0) | 0.09 |
| **TST** | **8.4 (7.9-8.8)** | **7.0 (6.5-7.5)** | **0.03** |
| C1 | 2.0 (1.0-3.0) | 2.0 (1.0-2.0) | 0.45 |
| C2 | 0.0 (0.0-0.0) | 1.0 (0.0-1.0) | 0.10 |
| C3 | 0.0 (0.0-1.0) | 1.0 (0.0-1.0) | 0.27 |
| C4 | 0.0 (0.0-0.0) | 0.0 (0.0-1.0) | 0.24 |
| C5 | 1.0 (1.0-1.0) | 1.0 (1.0-2.0) | 0.06 |
| C6 | 0.0 (0.0-0.0) | 0.0 (0.0-1.0) | 0.36 |
| C7 | 0.0 (0.0-1.0) | 1.0 (0.0-1.0) | 0.22 |
| SCOPA-AUT total | 13.0 (9.0-20.0) | 12.0 (6.0-19.0) | 0.95 |
| SCOPA-AUT Orthostatic hypotension § |  |  |  |
| No | 66.7% | 52.4% | 0.75 |
| Borderline | 22.2% | 42.9% | 0.51 |
| Yes | 11.1% | 4.8% | 1.00 |
| SCOPA-AUT Constipation § |  |  |  |
| No | 44.4% | 19.0% | 0.32 |
| Borderline | 22.2% | 57.1% | 0.17 |
| Yes | 33.3% | 23.8% | 0.93 |
| SCOPA-AUT Urinary § |  |  |  |
| No | 66.7% | 52.4% | 1.00 |
| Borderline | 22.2% | 42.9% |  |
| Yes | 11.1% | 4.8% |  |
| SCOPA-AUT Erectile Dysfunction § |  |  |  |
| No | 11.1% | 9.5% | 1.00 |
| Borderline | 22.2% | 23.8% | 1.00 |
| Yes | 22.2% | 23.8% | 1.00 |
| Female | 44.4% | 42.9% | 1.00 |
| MDS-UPDRS Part III | 0.0 (0.0-1.0) | 1.0 (0.0-3.0) | 0.20 |
| Action Tremor | 0.0 (0.0-0.0) | 0.0 (0.0-0.0) | 0.37 |
| Postural Tremor | 0.0 (0.0-0.0) | 0.0 (0.0-0.0) | 0.37 |
| Part III minus Tremor | 0.0 (0.0-1.0) | 0.0 (0.0-2.0) | 0.45 |
| Conversion time | 3.0 (2.0-4.0) | 3.0 (2.0-3.0) | 0.41 |

BMI, Body Mass Index; DEB, Dream enactment behavior; MMSE, Mini Mental State Examination; MoCA, Montreal Cognitive Assessment; PSQI, Pittsburgh Sleep Quality Index; RLS, Restless legs
syndrome; SCOPA-AUT, Scale for Outcomes in Parkinson's disease for Autonomic symptoms; K-GDS, Korean version of the Geriatric Depression Scale
\* Coffee use represents the average daily intake (cups per day), with Caffeine use defined as "yes" if Coffee use > 0.
\* Olfactory loss is defined as 1.5 standard deviation or less on a KVSS 1 or 2 test. KVSS (Korean Version of Sniffin' Sticks) is a standardized olfactory function test used to assess odor threshold, discrimination and identification.
† Coffee use represents the average daily intake (cups per day), with Caffeine use defined as "yes" if Coffee use > 0.
‡ Elementary school graduate or lacking formal education.
§ For each SCOPA-AUT subcategory, the subcategory is categorized as Yes if there is a score of 2 or more in that corresponding items; No if all are 0; otherwise, Borderline.

**Table 3.**
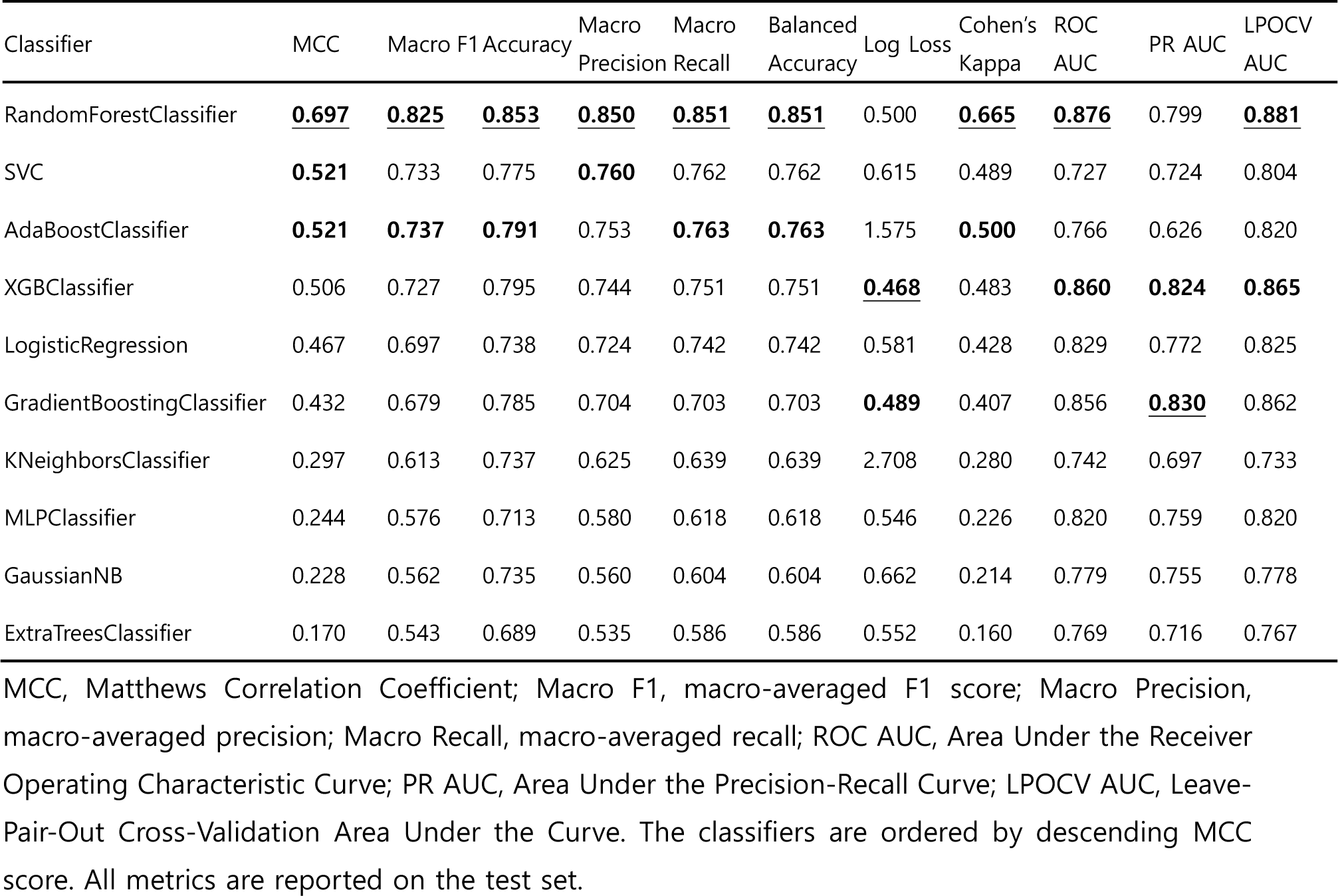
Performance of Classifiers developed with the mRMR feature set.

The features selected by RFE-RSF included weight, UPDRS III excluding tremor (the Unified Parkinson’s Disease Rating Scale score excluding tremor components), antidepressant use, RBDQ-KR factor 2 (capturing behavioral aspects from the REM Sleep Behavior Disorder Questionnaire), coffee use, and age.

SHAP analysis provided valuable insights into the relationships between features and phenoconversion risk. Features such as Age, Antidepressant use, UPDRS III excluding tremor, and Coffee use, which were significant in univariable analysis, showed linear relationships with phenoconversion risk (Supplementary Figure 7). An increase in values for Age, Antidepressant use, and UPDRS III excluding tremor corresponded to a higher risk of phenoconversion. Conversely, higher Coffee use, a protective factor, was associated with lower SHAP values, indicating a reduction in risk. Body weight and RBDQ-KR factor 2 were not identified as significant factors in the univariable analysis. When analyzing SHAP values, they appeared to have a non-linear pattern with phenoconversion risk. The risk associated with Weight was elevated in the 55-65kg range. RBDQ-KR Factor 2 displayed a complex pattern, with increased risk at scores below 20 and between 35-45. When evaluating the mean absolute SHAP values, which indicate the overall impact of each feature on the model’s output, Age was found to be the most influential factor. This was followed, in order of importance, by RBDQ-KR factor 2, Weight, Antidepressant use, Coffee use, and UPDRS III excluding tremor. (Figure 3) These results highlight the significant role of age as a predictor in the model and the varying degrees of influence of other factors.

**Figure 3.**
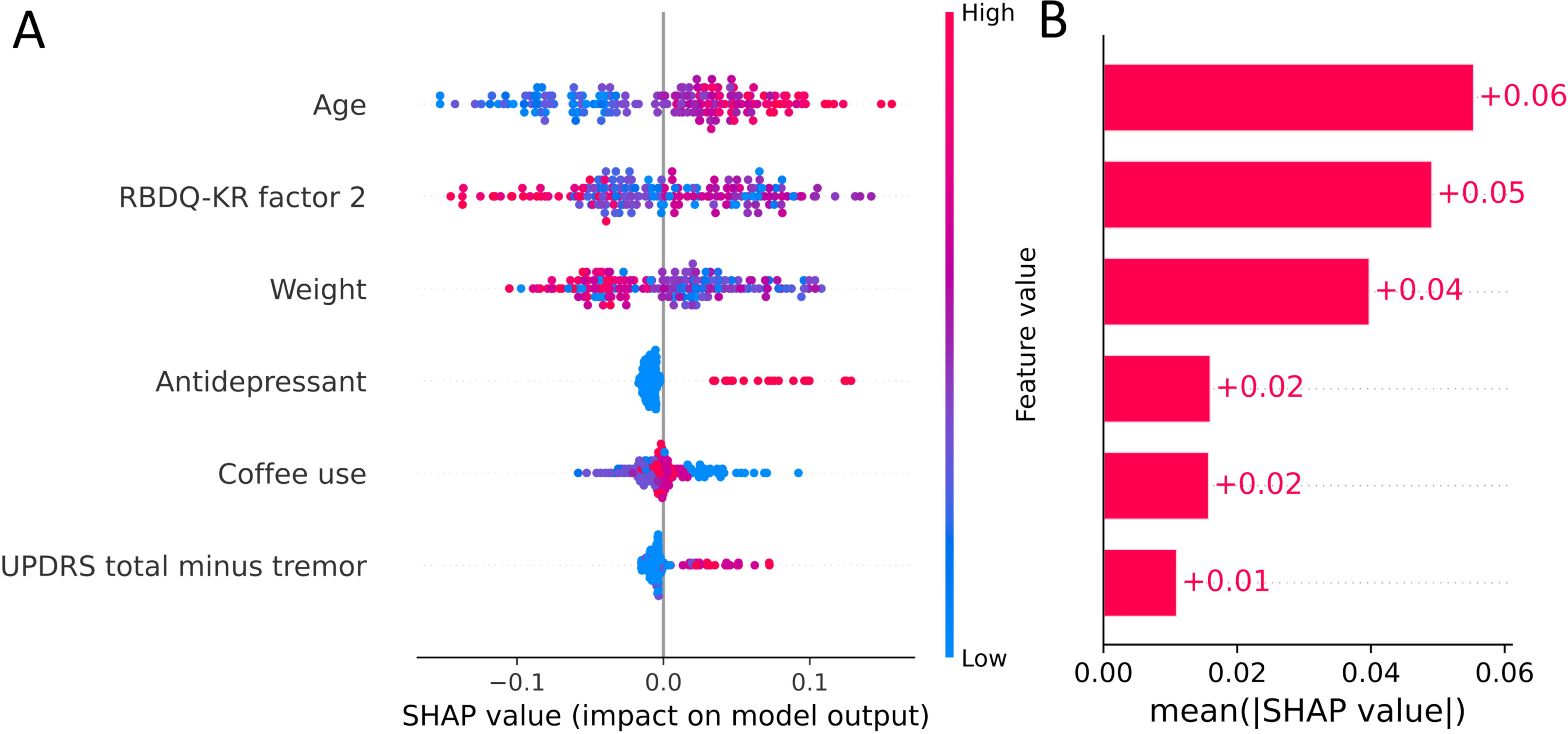
Two Plots for a Time Prediction Model: (A) Beeswarm Plot of SHAP Values for Each Feature (B) Bar Plot of Mean Absolute SHAP Values, Sorting Features by Importance

### Phenoconversion Subtype Prediction

In addition to developing a model for predicting the time to phenoconversion, we also focused on distinguishing the subtype of phenoconversion—specifically, whether patients would exhibit a motor-first or cognition-first progression. We identified several clinical indicators that were significantly different between motor-first and cognition-first in a univariable analysis, including age, Mini-Mental State Examination (MMSE), Montreal Cognitive Assessment (MoCA), and the Pittsburgh Sleep Quality Index-Total Sleep Time (PSQI-TST). In assessing MIC values, PSQI-TST and MMSE showed significantly high contributions (Supplementary Table 2). We then utilized the mRMR method for optimal feature selection resulting in PSQI-TST, MoCA, and age. Additionally, the Boruta algorithm was implemented, affirming the same set of features as identified by mRMR.

We developed ten classifiers to predict phenoconversion subtypes, utilizing univariable feature set and mRMR feature set, and tried enhancing their performance with SMOTE and Self-Training. Among these, the RandomForestClassifier (RF) showed the best performance across both feature sets, particularly with mRMR, achieving the highest MCC value of 0.697 through 100 repeated 5-fold stratified cross-validation (Supplementary Figure 4, and 5). RF also excelled in metrics like Macro F1, Accuracy, Precision, Recall, Balanced Accuracy, Cohen’s Kappa, and LPOCV AUC, demonstrating its robustness.

When using RF and mRFR feature sets to get SHAP values, PSQI-TST was the most important, followed by MoCA and Age (Supplementary Figure 6). PSQI-TST of less than 8 hours contributed to prediction to motor-first, MoCA of 25 or more and Age of 70 or less contributed to prediction to motor-first (Supplementary Figure 8).

### Web Deployment

A user-friendly web application was developed for physician access, which included both models: the XGBSE-KN model for phenoconversion time and the RF model for subtype prediction. The application offers individualized predictions and rationales based on input characteristics, aiding physicians in making informed decisions about patient management and prognosis. (Supplementary Figure 9). To demonstrate the use case of the model, we generated a report using actual patient cases, which is shown in Figure 4.

**Figure 4.**
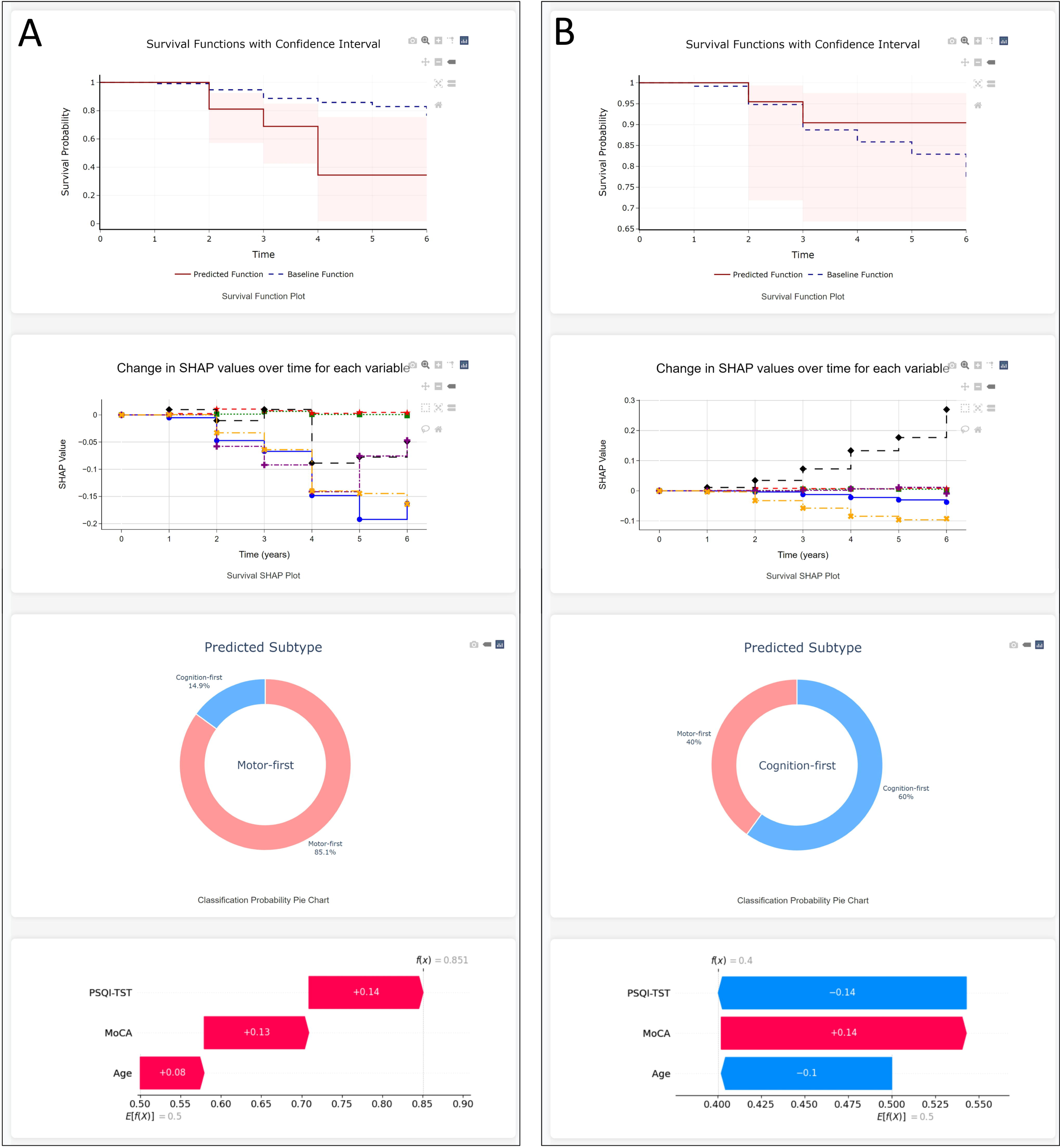
Illustrative Examples of Reports Utilizing Patient Data. (A) This report illustrates the case of a patient in their 70s with a seven-year history of dream enactment behaviors. Initially presenting with an MDS-UPDRS-III score of 0 and a MoCA-K score in the normal range, the patient also showed signs of mild depression (HADS-depression score of 9, GDS-K of 14). Over a period of 3.9 years, the patient developed Parkinson’s disease. The model’s predicted survival function suggests a progressive decline in survival probability free of phenoconversion, dropping significantly by the fourth year. SHAP analysis identifies the patient’s age, lack of antidepressant use, and other factors as influential in the patient’s rapid phenoconversion to a motor-first subtype, which is supported by the actual clinical outcome. The SHAP analysis, factoring in age, MoCA score, and PSQI-TST score, reinforces the motor-first subtype prediction. (B) This report concerns a patient in their 70s who developed dream enactment behavior three years before the RBD diagnosis. The patient presented with an initial MDS-UPDRS III score of 0 and a MoCA-K score in the mild cognitive impairment range, alongside a decreased sense of smell as indicated by a KVSS2 score of 16. After 6.2 years, the patient remains free from neurodegenerative disease. The model’s survival prediction suggests a high likelihood of remaining phenoconversion-free (90% at 6 years). SurvSHAP analysis denotes the patient’s age as a risk factor and RBDQ-KR Factor 2 as protective against phenoconversion. The model’s subtype prediction leans towards a cognition-first manifestation. SHAP analysis identifies the patient’s age and PSQI-TST score as significant factors in this subtype prediction.

## Discussion

In this study, we developed machine learning models to predict phenoconversion time and subtype in patients with iRBD. Leveraging a relatively large patient cohort with comprehensive clinical markers, our findings provided important insights into phenoconversion predictors, and increased confidence in the generated model.

The XGBSE-KN model, using features selected through the RFE-RSF method, was the most effective model for phenoconversion time prediction, achieving an excellent concordance index of 0.823 and a good Integrated Brier Score (IBS) of 0.123. For subtype classification, the RF demonstrated the highest performance, with a good to very good Matthews Correlation Coefficient (MCC) of 0.697. These results underscore the potential of advanced machine learning techniques in accurately predicting the prognosis in patients with RBD.

Our research encompassed a cohort of 178 patients with iRBD, utilizing an extensive array of clinical variables to enhance the study’s reliability and value. Over a median follow-up duration of 3.6 years, 30 individuals progressed to a neurodegenerative condition. Our study identified several critical predictors of phenoconversion, including age, antidepressant use, solvent exposure, and caffeine consumption. Additionally, components of the PSQI — PSQI-C3 (sleep duration) and PSQI-C4 (sleep efficiency), and MDS-UPDRS part III, excluding tremor scores, were found to be significant.

Old age was identified as a significant predictor, which aligns with existing research suggesting a higher vulnerability to neurodegenerative diseases as one ages. Solvent exposure is a known risk factor for Parkinson’s disease. The coffee is also known for protective effect against Parkinson’s disease. In our cohort, solvent exposure increased the risk of phenoconversion, and coffee consumption had a protective relationship with phenoconversion. This was parallel to that observed in Parkinson’s disease. In our study, we also found that antidepressant use at the baseline evaluation was a significant risk factor for phenoconversion in patients with Rapid Eye Movement (REM) Sleep Behavior Disorder (RBD). Antidepressant use has been associated with symptoms of RBD. Patients with iRBD who take antidepressants show significant abnormalities in neurodegenerative markers, such as olfaction, color vision, and motor function. A previous study using 18F-DOPA showed that 18F-DOPA uptake was significantly lower in patients with co-morbid RBD and MDD compared to patients taking medication for MDD. It suggests that the development of RBD symptoms in MDD patients is not simply an antidepressant-induced condition, but an early stage of synucleinopathic neurodegeneration. ^31^

Selected features for the final model were slightly different from the features identified with statistical methods. Age, Antidepressant use, UPDRS III excluding tremor, and Coffee use were identified in univariable analysis and the SHAP analysis revealed the linear relationship with the phenoconversion risk. On the other hand, RBDQ-KR Factor 2, Weight were not identified with the statistical methods and SHAP analysis revealed non-linear relationships with the phenoconversion risk. This illustrates the capability of machine learning models to uncover intricate relationships that traditional statistical methods might overlook, especially in large variable sets. It might be possible to identify these non-linear relationship by using strategies like stratifying the variables, but it would not be easy to find these relationships among many variables without missing them.

Our approach also addresses the limitations of traditional survival analysis methods like CoxPH. Many studies with survival analyses use the CoxPH model for its solid theoretical basis.^32^ However, its effectiveness is constrained by its reliance on linear feature interactions and its incompatibility with data exhibiting multicollinearity or possessing large-scale, high-dimensional feature sets.

In examining the subtypes of phenoconversion, Cognition-first patients had lower baseline MMSE and MoCA scores, which was confirmed by previous studies.^5^ The fact that the Total Sleep Time (TST) component of the Pittsburgh Sleep Quality Index (PSQI-TST) was longer in the cognition-first group is an intriguing finding. Typically, cognitive decline is often associated with sleep disturbances or reduced total sleep time. A study comparing PSG findings in PD and DLB found that DLB had lower sleep efficiency, total sleep time, and REM sleep duration, and higher sleep latency and WASO.^33^ This longer PSQI-TST in the cognition-first group might indicate a compensatory mechanism where the body requires more rest due to cognitive stresses, or it could be reflective of a less efficient sleep, where despite longer durations, the quality or the restorative aspects of sleep are compromised. In Addition, although PSQI-TST is typically used to measure the actual time spent sleeping, it is possible that patients might not exclude periods of WASO, thereby reporting longer sleep durations in the PSQI-TST, even though their effective sleep time might be reduced due to frequent awakenings or disturbances. This is supported by a previous large study comparing polysomnography, actigraphy, and sleep diaries, which found that sleep diaries tend to understate sleep latency and WASO, and exaggerate TIB, TST, and SE.^34^

We found that the cognition-first group was older than the motor-first group at the baseline. Aging is a significant risk factor for cognitive decline. The older population is naturally more susceptible to cognitive impairments due to various factors, including comorbidities, reduced cognitive reserve, and age-related changes in brain structure and function. Therefore, older patients with iRBD may be more likely to manifest cognitive symptoms first due to their greater baseline vulnerability to cognitive decline.

To capture non-linear relationships between subtypes and variables, we utilized MIC. However, in subtype classification, it was hard to find such non-linear features. Small sample sizes may have hindered the ability to find these factors, or perhaps none of the clinical metrics we collected have a non-linear relationship.

Another critical aspect of this study was the development of a user-friendly web application for physicians, facilitating the application of these complex machine learning models in a clinical setting. This approach underscores the potential of machine learning in providing more accurate and personalized insights in medical practice.

## Limitations

Our study has some limitations. One of the limitations of this study is that it was conducted at a single center without an external validation data. While this approach enables consistent protocols and data collection, it reduces the generalizability of the findings. Additionally, the sample size, while relatively large for a single-center study, is relatively small, which may also reduce generalizability. This limitation is particularly evident in our subtype prediction model, where only 30 eligible patients were included, and merely 3 features were selected, challenging the achievement of higher accuracy. In the field of machine learning, larger datasets may provide more robust and generalizable findings. A limited sample size can restrict the model’s ability to capture more complex or subtle relationships within the data. Given a small sample size and a large number of predictors, there is an inherent potential of overfitting the models. As a consequence, the representativeness of the model and findings may be confined to similar environments or populations. While techniques such as cross-validation are used to mitigate this risk, it remains a concern in studies with smaller datasets.

Another limitation is the relatively short follow-up period. Since neurodegenerative diseases often progress slowly, a longer follow-up duration would provide more comprehensive data on the phenoconversion process and the progression of symptoms over an extended period.

Although we collected comprehensive data, the scope of data collected may not encompass all potential factors influencing phenoconversion in iRBD. There are numerous variables, both known and unknown, that could play a role in the progression of neurodegenerative diseases in patients with iRBD. Excluding certain environmental, genetic, or lifestyle factors not captured in the study might have led to an incomplete picture of the phenoconversion process.

These limitations highlight the need for further research in more diverse and larger populations, possibly through multicenter studies. Expanding data collection and using larger datasets could improve understanding of iRBD phenoconversion. Additionally, it would be necessary to validate the predictive models in external cohorts to assess their generalizability and robustness in different settings.

## Conclusions

This study has developed a practical and accurate predictive model for iRBD patients, providing valuable insights for clinical treatment planning and prognosis. It highlights the significant role of machine learning in enhancing the precision and personalization of medical care, particularly in the context of predicting phenoconversion in iRBD patients.

## Acknowledgments

This work was supported by the Brain Research Program through the National Research Foundation of Korea (NRF) funded by the Ministry of Science, ICT & Future Planning (2017M3C7A1029688) and the National Research Foundation of Korea (NRF) grant funded by the Korean government (MSIP) (2017R1A2B2012280).

## Disclosure Statement

Financial Disclosure: none

Non-financial Disclosure: none

## Data availability Statement

The data that support the findings of this study are available from the corresponding author upon reasonable request.

## Notes

### Competing Interest Statement

The authors have declared no competing interest.

### Author Declarations

The study was approved by the Institutional Review Board of Seoul National University Hospital (IRB No.: 1708-169-883).

