## Supplemental Tables for "FORECAST-RBD: Forecasting Phenoconversion Risks and Its Clinical Phenotype in Patients with Isolated RBD Using Machine Learning and Explainable AI"

### Supplementary Table 1. Features selected by different feature selection methods in survival analysis.

| Method | Selected Features |
| --- | --- |
| Univariable | Age, Antidepressant, Caffeine use, Education, MMSE, MoCA, PSQI-C3, PSQI-TST, Solvent exposure, UPDRS III, UPDRS III excluding tremor |
| Lasso | Age, Antidepressant, UPDRS III excluding tremor |
| RFE-RSF | Age, Antidepressant, Coffee use, RBDQ-KR factor 2, UPDRS III excluding tremor, Weight |
| RFE-GBS | Age, Antidepressant, BMI, Coffee use, K-GDS, PSQI-C4, PSQI-TST, RBDQ-KR factor 2, UPDRS III excluding tremor, Weight |
| SKB-RSF | Age, Antidepressant, Caffeine use, Coffee use, MoCA, RBDQ-KR factor 2, Solvent exposure |
| SKB-GBS | Age, Antidepressant, Coffee use, RBDQ-KR Factor 2 |

K-GDS, Korean version of the Geriatric Depression Scale; RFE, Recursive Feature Elimination; RSF, Random Survival Forest; GBS, Gradient Boosting Survival Analysis; MMSE, Mini-Mental State Examination; MoCA, Montreal Cognitive Assessment; PSQI-C3, A component of the Pittsburgh Sleep Quality Index related to sleep duration; PSQI-TST, Total Sleep Time in the Pittsburgh Sleep Quality Index; UPDRS III, Unified Parkinson's Disease Rating Scale Part III; UPDRS III excluding tremor, A metric within UPDRS III excluding tremor assessment; RBDQ-KR factor 2, A factor within the REM Sleep Behavior Disorder Questionnaire - Korea version, assessing behavioral symptoms.

Note: Coffee use represents the average daily intake (cups per day), with Caffeine use defined as "yes" if Coffee use > 0.

### Supplementary Table 2. Maximal Information Coefficient (MIC) Values for Differentiating Motor-first and Cognition-first Groups

| Variable | MIC | P-value |
| --- | --- | --- |
| **PSQI-TST** | **0.435689** | **0.00570** |
| **MMSE** | **0.293764** | **0.02770** |
| MoCA | 0.281291 | 0.05630 |
| Age | 0.256128 | 0.14465 |
| RBDQ-KR Factor 2 | 0.193507 | 0.38760 |
| ISI | 0.187807 | 0.44660 |
| GDS-K | 0.186189 | 0.42890 |
| PSQI-Total | 0.174716 | 0.41690 |
| Weight | 0.160395 | 0.68530 |

MIC, Maximal Information Coefficient; PSQI-TST, Pittsburgh Sleep Quality Index - Total Sleep Time; MMSE, Mini-Mental State Examination; MoCA, Montreal Cognitive Assessment; RBDQ-KR Factor 2, REM Sleep Behavior Disorder Questionnaire-Korea, Factor 2; ISI, Insomnia Severity Index; GDS-K, Geriatric Depression Scale-Korean version; PSQI-Total, Pittsburgh Sleep Quality Index Total Score.

### Supplementary Table 3. Hyperparameters for the XGBSE-KaplanNeighbor Survival Model

| Parameter | | Value | Description |
| --- | --- | --- | --- |
| n_neighbors | | 24 | Number of neighbors for computing KM estimates |
| radius | | None | The radius limit for the neighborhood search. None indicates no limit. |
| xgb_params: | |  | Parameters for the XGBoost component of the model. |
|  | booster | dart | The type of booster used in the XGBoost model |
|  | colsample_bynode | 0.794139987 | The fraction of columns to be subsampled for each node. |
|  | eval_metric | cox-nloglik | Evaluation metrics for validation data, set to negative partial log-likelihood for Cox proportional hazards regression |
|  | learning_rate | 0.026334112 | The step size at which the model's weights are updated during training. |
|  | max_depth | 59 | The maximum depth of the decision trees in the XGBoost model. |
|  | min_child_weight | 0.736347122 | The minimum sum of instance weight needed in a child node. |
|  | objective | survival:cox | The objective function for the XGBoost model, set to Cox proportional hazards. |
|  | subsample | 0.893111458 | Subsample ratio of the training instances. |
|  | tree_method | exact | The tree construction algorithm used in XGBoost |

### Supplementary Table 4. Hyperparameters for the RandomForestClassifier

| **Parameter** | **Value** | **Description** |
| --- | --- | --- |
| bootstrap | FALSE | Whether bootstrap samples are used when building trees. |
| ccp_alpha | 0 | Complexity parameter used for Minimal Cost-Complexity Pruning. |
| class_weight | Balanced | The “balanced” mode uses the values of y to automatically adjust weights inversely proportional to class frequencies in the input data |
| criterion | gini | The function to measure the quality of a split. |
| max_depth | 50 | The maximum depth of the tree. |
| max_features | 1 | The number of features to consider when looking for the best split. |
| max_leaf_nodes | None | Grow trees with max_leaf_nodes in best-first fashion. |
| max_samples | None | If bootstrap is True, the number of samples to draw from X to train each base estimator. |
| min_impurity_decrease | 0 | A node will be split if this split induces a decrease of the impurity greater than or equal to this value. |
| min_samples_leaf | 8 | The minimum number of samples required to be at a leaf node. |
| min_samples_split | 7 | The minimum number of samples required to split an internal node. |
| min_weight_fraction_leaf | 0 | The minimum weighted fraction of the sum total of weights (of all the input samples) required to be at a leaf node. |
| monotonic_cst | None | Constraint to enforce monotonic relationship between features and target. If monotonic_cst is None, no constraints are applied. |
| n_estimators | 269 | The number of trees in the forest. |
| oob_score | FALSE | Whether to use out-of-bag samples to estimate the generalization score. |
