## Supplemental Figures for "FORECAST-RBD: Forecasting Phenoconversion Risks and Its Clinical Phenotype in Patients with Isolated RBD Using Machine Learning and Explainable AI"

### Supplementary Figure 1. Overview of the study design and methodology

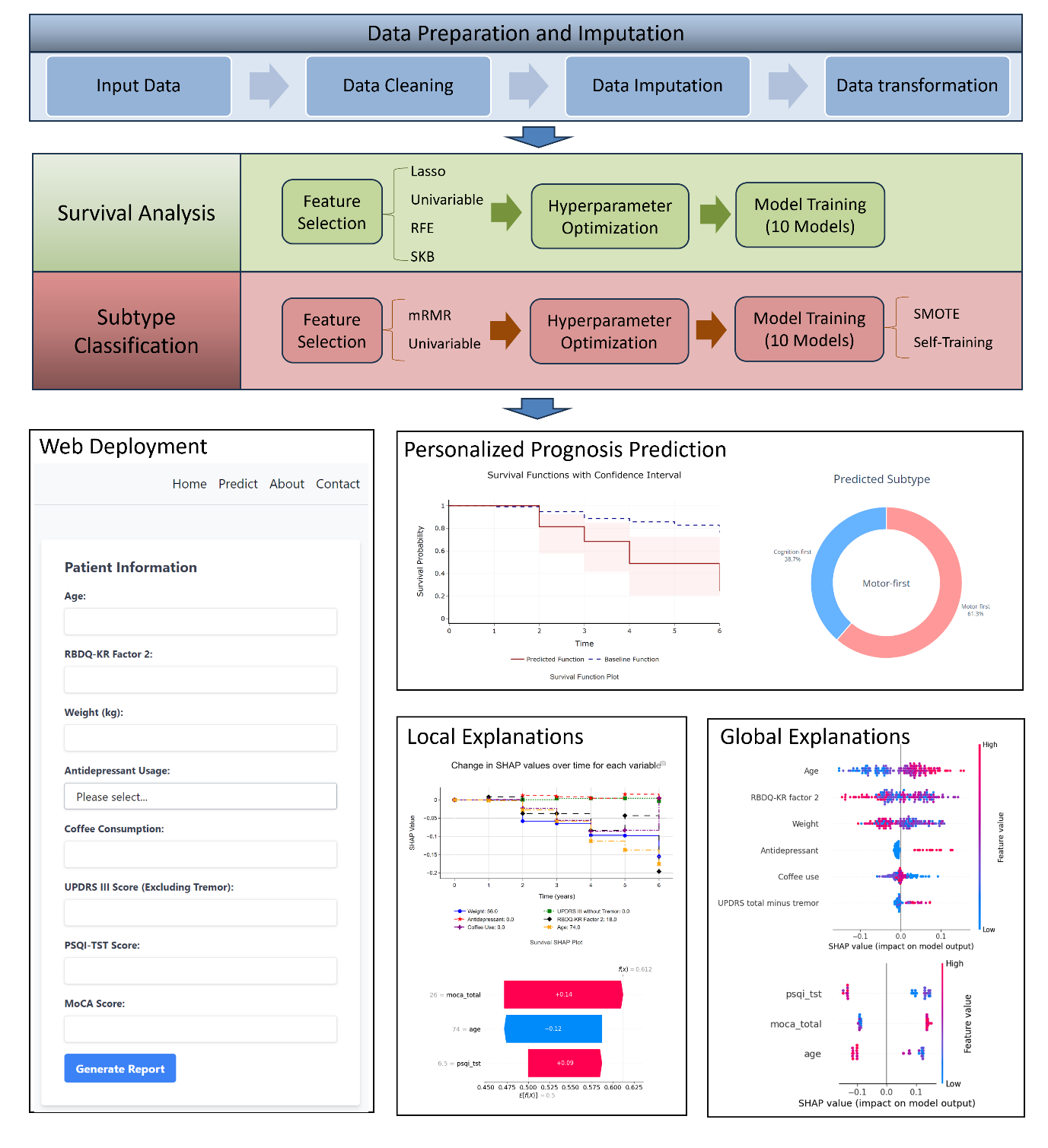

### Supplementary Figure 2. Evaluation of phenoconversion time prediction models using different feature selection methods based on Uno's Concordance Index

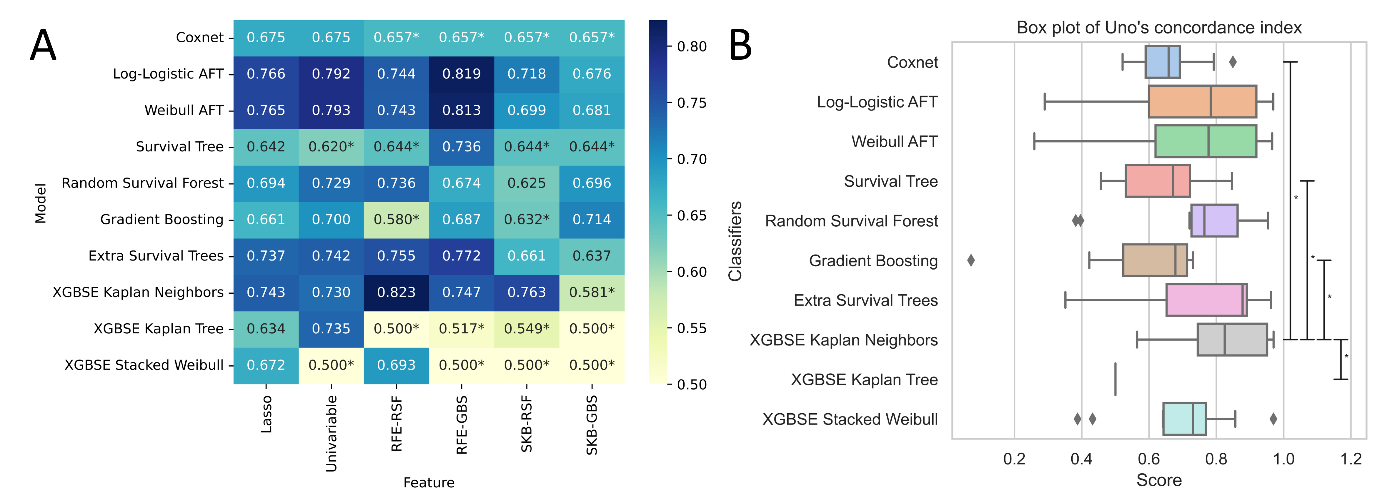
 (A) Uno's concordance index values by model and feature selection method. (B) Uno’s concordance index values for models utilizing the RFE-RSF (Recursive Feature Elimination-Random Survival Forests) feature set.

### Supplementary Figure 3. Evaluation of phenoconversion time prediction models using different feature selection methods based on the Integrated Brier Score.

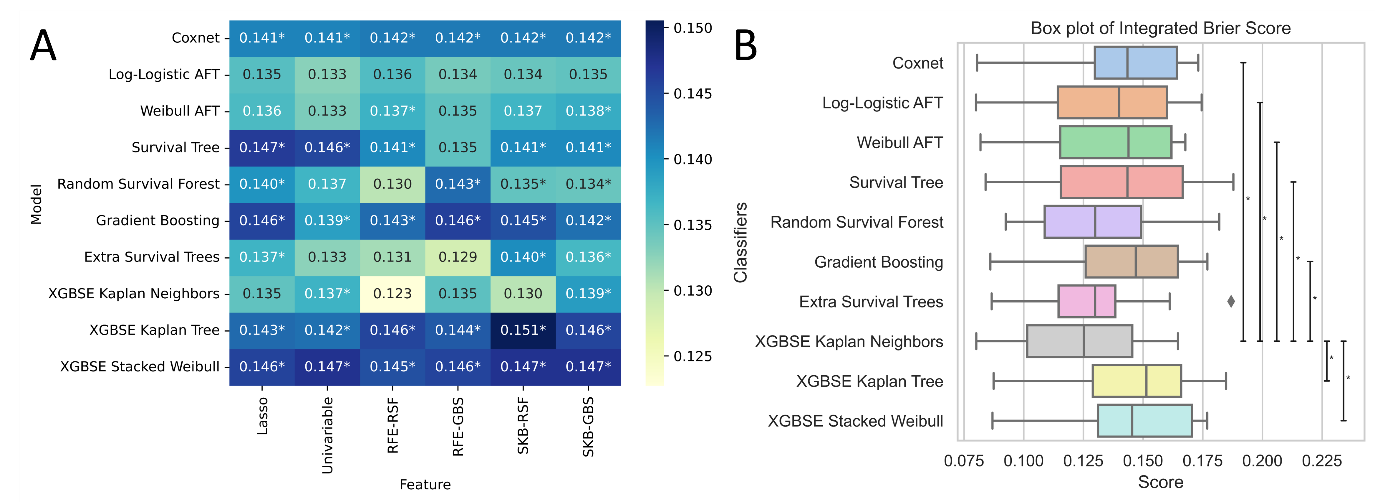

(A) Integrated Brier scores by model and feature selection method. (B) Integrated Brier scores of models using the RFE-RSF (Recursive Feature Elimination-Random Survival Forests) feature set.

### Supplementary Figure 4. Performance of classifiers for phenoconversion subtype prediction using the minimum-redundancy-maximum-relevance (mRMR) feature set.

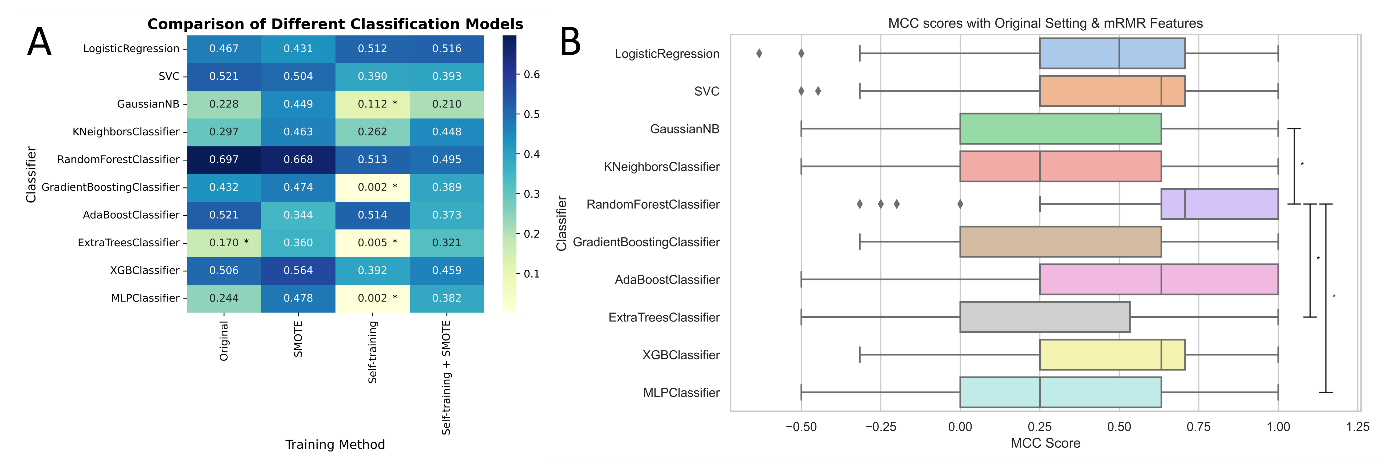

(A) Matthews Correlation Coefficient scores of classifiers using mRMR features. (B) Matthews Correlation Coefficient scores of models using original settings and the mRMR feature set.

### Supplementary Figure 5. Evaluation of classifiers for phenoconversion subtype prediction using univariable features.

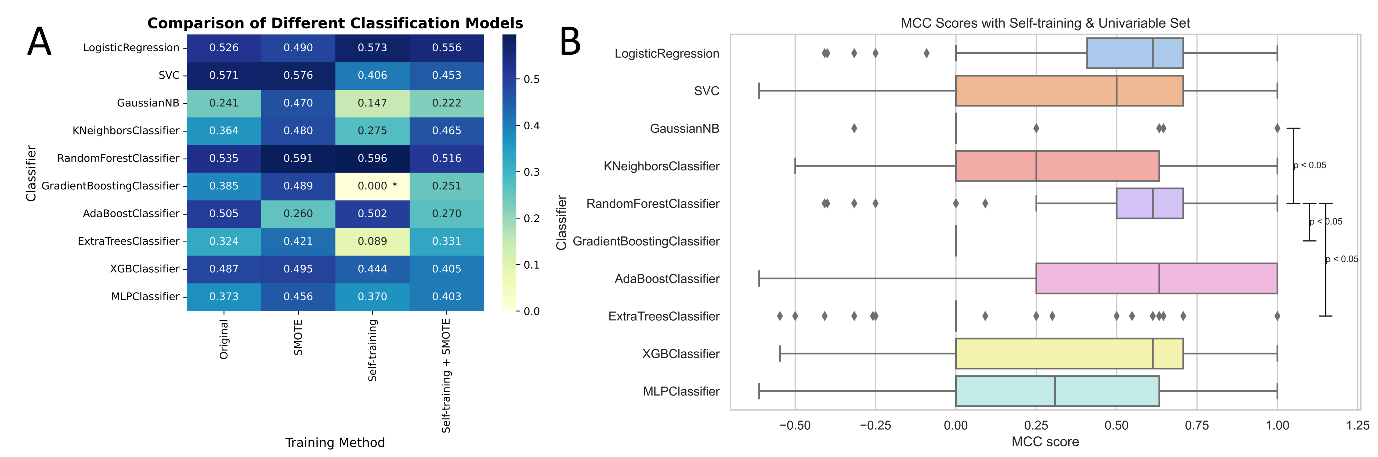

(A) MCC scores of classifiers using univariable features. (B) MCC scores of models using Self-training and the univariable feature set.

### Supplementary Figure 6. Explanation of the phenoconversion subtype prediction model using SHapley Additive exPlanations (SHAP).

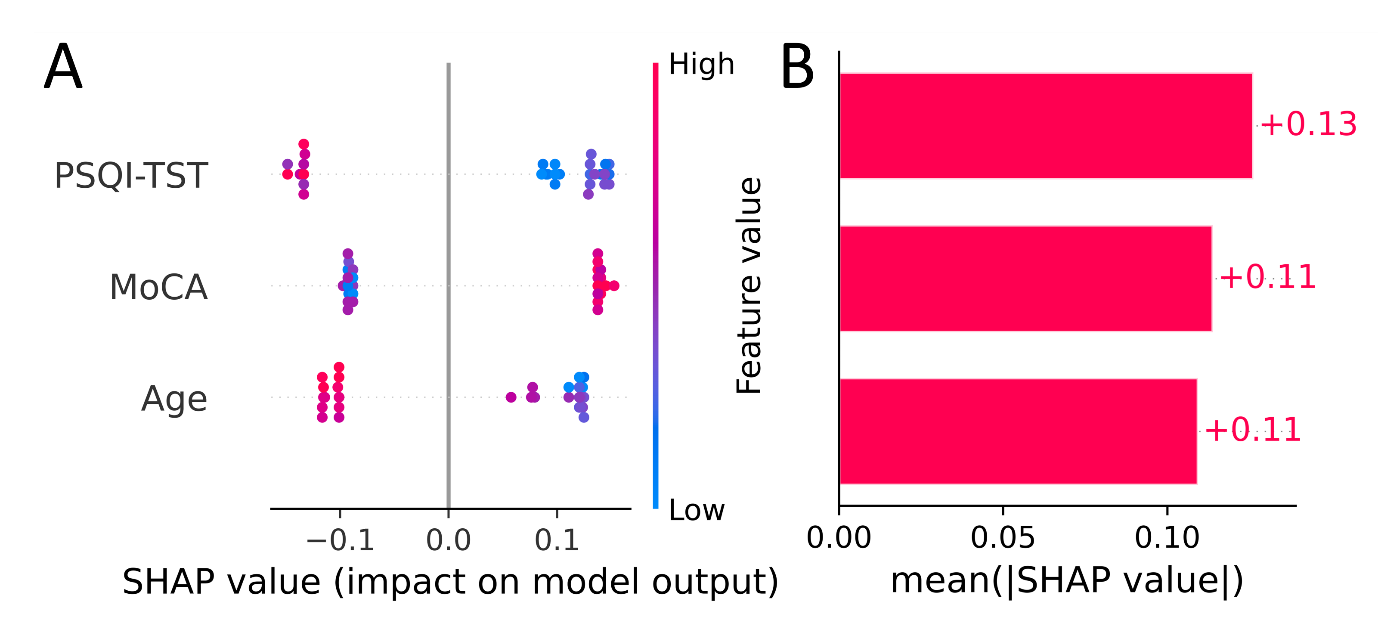
(A) Beeswarm plot of SHAP values for each feature. (B) Bar plot of mean absolute SHAP values, sorting features by importance.

### Supplementary Figure 7. SHAP dependence plots for selected features in the phenoconversion time prediction model.

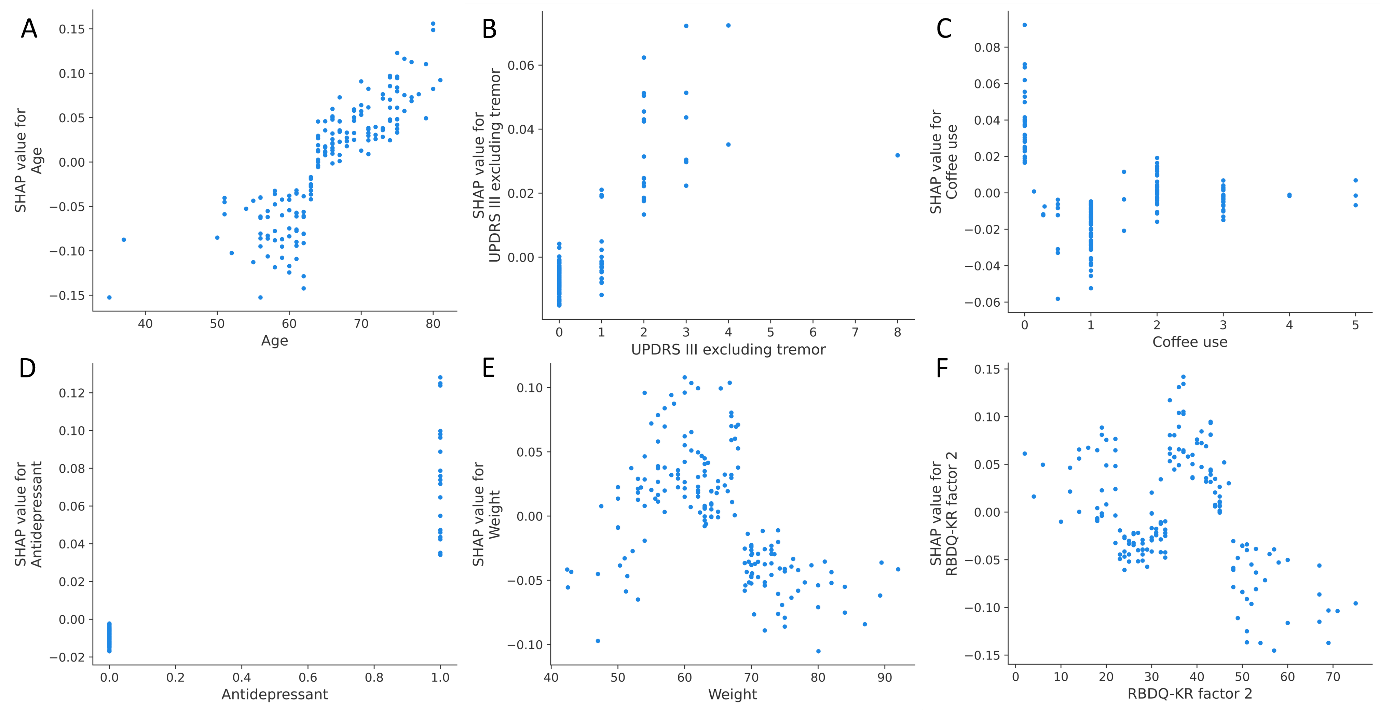

(A) Age. (B) Unified Parkinson's Disease Rating Scale (UPDRS) Part III excluding tremor. (C) Coffee use. (D) Antidepressant use. (E) Weight. (F) REM Sleep Behavior Disorder Questionnaire-Korea (RBDQ-KR) factor 2.

### Supplementary Figure 8. SHAP dependence plots for selected features in the phenoconversion subtype prediction model.

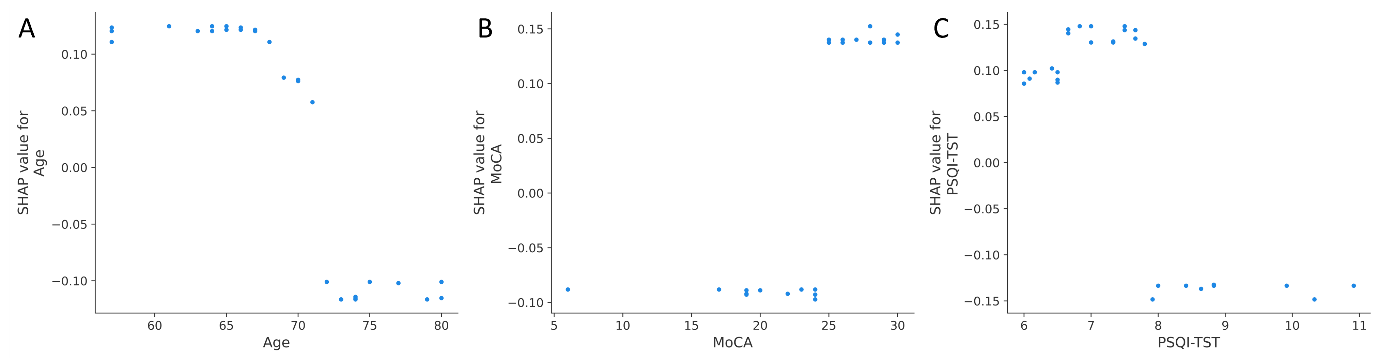
(A) Age. (B) Montreal Cognitive Assessment (MoCA) score. (C) Pittsburgh Sleep Quality Index-Total Sleep Time (PSQI-TST).

### Supplementary Figure 9. Web application for phenoconversion prediction in isolated REM sleep behavior disorder (iRBD).

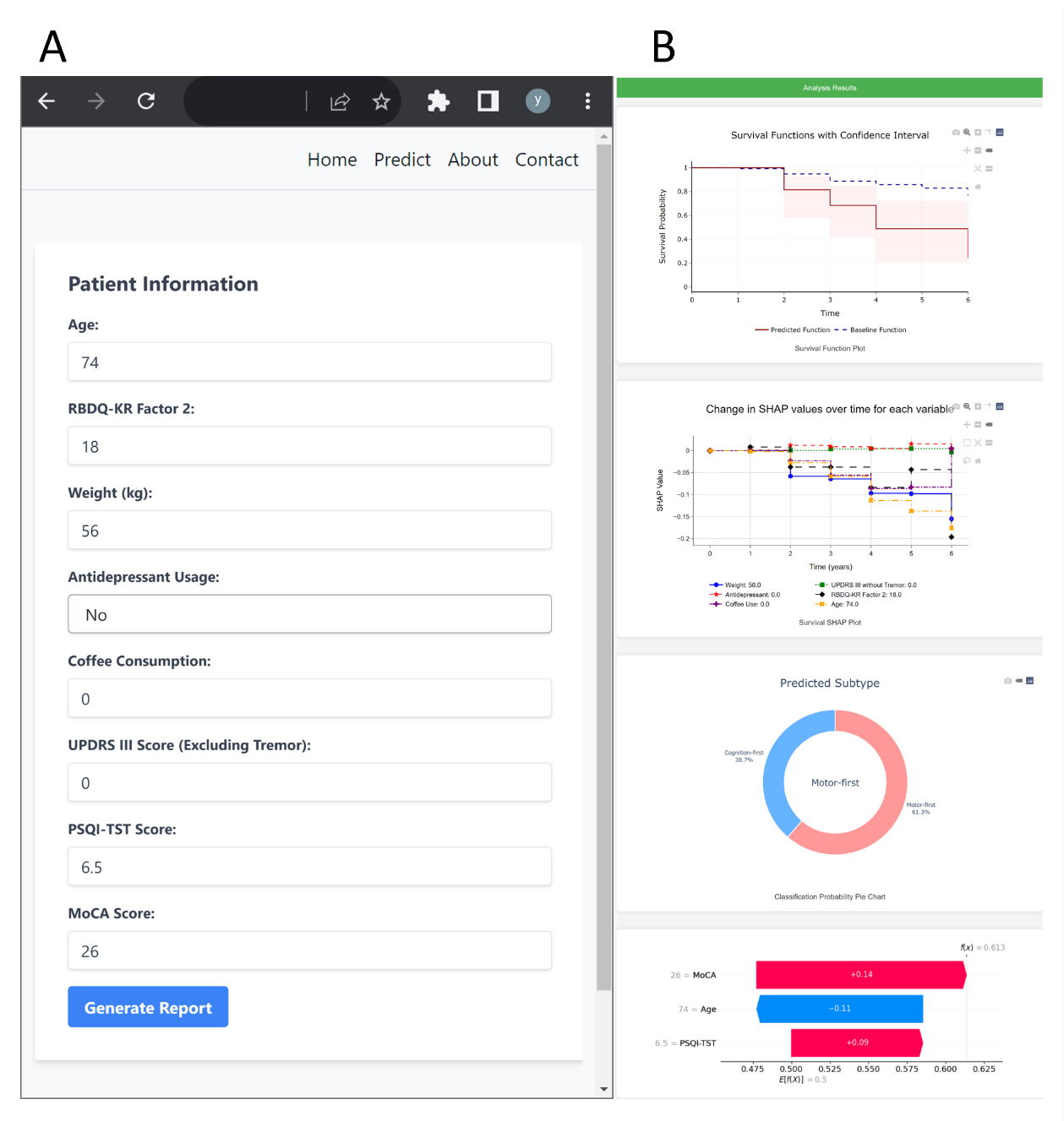

(A) Clinical indicator input page. (B) Example of a generated report based on the input data.
